# Parental preferences for a mandatory vaccination scheme in England; a discrete choice experiment

**DOI:** 10.1101/2021.12.22.21268231

**Authors:** Louise E Smith, Ben Carter

**Author notes:** Corresponding author: Louise E Smith, Post-doctoral Researcher. Department of Psychological Medicine, King’s College London, Weston Education Centre, Cutcombe Road, London, SE5 9RJ. Twitter handle: @louisesmith142, Ben Carter, Reader in Medical Statistics. Biostatistics and Health Informatics, King’s College London, 16 De Crespigny Park, London SE5 8AB. Twitter handle: @DrBenCarter. Both authors contributed equally to the work.

## Abstract

**Background:** Mandatory vaccination has been mooted to combat falling childhood vaccine uptake rates in England. This study investigated parental preferences for a mandatory vaccination scheme.

**Methods:** Discrete choice experiment. Six attributes were investigated: vaccine (MMR, 6-in-1), child age group (2 years and older, 5 years and older), incentive (£130 cash incentive for parent, £130 voucher incentive for child, no incentive), penalty (£450 fine, parent not able to claim Child Benefits for an unvaccinated child, unvaccinated child not able to attend school or day care), ability to opt out (medical exemption only, medical and religious belief exemption), and compensation scheme (not offered, offered). Mixed effects conditional logit regression models were used to investigate parental preferences and relative importance of attributes.

**Findings:** Participants were 1,001 parents of children aged 5 years and under (53% female, mean age=33·6 years, SD=7·1, 84% white British). Parental preferences were mostly based on incentives (slight preference for cash pay-out for the parent versus a voucher for the child) and penalties (preference for schemes that did not allow unvaccinated children to attend school or day care and those that withheld financial benefits for parents of unvaccinated children). Parents also preferred schemes that: offered a compensation scheme, mandated the 6-in-1 vaccine, mandated vaccination in children aged 2 years and older, and that offered only medical exemptions.

**Interpretation:** Results can inform policymakers’ decisions about how best to implement a mandatory childhood vaccination scheme in England.

**Funding:** Data collection was funded by a British Academy/Leverhulme Small Research Grants (SRG1920\101118).

**Research in context:** *Evidence before this study:* Uptake of childhood vaccines in high-income countries has decreased in recent years. Making vaccination mandatory has the potential to increase uptake. There is no standard approach to mandatory vaccination schemes. Research suggests that the cultural context will affect perceived acceptability of vaccine laws. Mandatory vaccination has been mooted in England as a way to increase vaccine uptake. However, there is no recent research investigating parental preferences for how a mandatory vaccine scheme could be implemented.

*Added value of this study:* We used a discrete choice experiment to investigate English parents’ preferences for a mandatory vaccination scheme. Variables included were parameters that are likely to be considered by policymakers if a mandatory vaccination scheme were to be proposed.

*Implications of all the available evidence:* Study results indicate that parents prefer mandatory vaccination schemes that offer financial incentives for vaccination. The penalty imposed for missing a vaccine dose, and the inclusion of a compensation scheme for severe adverse effects also influenced preferences. These results can be used to inform policy should a mandatory vaccination scheme be proposed in England.

## Introduction

The World Health Organization estimates that 4-5 million lives are saved by vaccination annually; a further 1.5 million deaths could be avoided if vaccine uptake improves.^1^ The COVID-19 pandemic has highlighted the importance of vaccination. However, uptake of childhood vaccines is declining in high-income countries.^2,3^ Where vaccinations have been mandated, recent rates of non-medical exemptions have increased.^4^ Currently, child vaccination is voluntary in the UK, but mandating it has been mooted to combat falling vaccine rates.^5^

There is no standard approach to mandatory vaccination schemes. Approaches vary by which vaccines are mandatory, age groups included, and flexibility of the mandate (e.g. penalties, enforcement, ability to opt out, compensation for serious adverse events). Of the 105 countries that mandated vaccination in December 2018, the most common penalty used was limiting the unvaccinated child’s access to schooling or day care.^6^ A recent review of vaccination laws in Europe found much variation between countries, and that no common “best approach” could be pinpointed.^7^ Instead, the report found that context in individual countries should be considered when drafting vaccine laws.

In England, there is a lack of research investigating parents’ views on, and preferences for, a mandatory vaccination scheme. This is partly because child vaccination has always been voluntary. The COVID-19 pandemic has re-ignited the debate around mandatory vaccination, with COVID-19 vaccination being mandated in frontline health and social care workers in England.^8^ Research into the acceptability of financial incentives and quasi-mandatory childhood vaccination schemes (where parents can opt their child out of vaccination for medical, religious, or philosophical reasons) in England indicates that parents prefer universal, rather than targeted, schemes, and that financial incentives are not considered appropriate motivation for vaccination.^9,10^ Quantitative research indicates that parents at high risk of incomplete vaccination prefer schemes offering cash incentives to no incentive; there was no preference in parents not at high risk of vaccine refusal.^9-11^ However, this study did not investigate parental preferences for other aspects of mandatory vaccination schemes that would need to be considered before implementation, such as the ability to opt out.

The aim of this study was to assess parental preferences for a mandatory vaccination scheme in parents of children aged five years and under who lived in England. Subgroup analyses assessed variation in preferences in groups identified *a-priori* as being more likely to refuse child vaccinations and less likely to approve of mandatory vaccination.

## Methods

Discrete choice experiments (DCE) allow researchers to investigate participant preferences for a number of pre-selected attributes, each with multiple levels.^12^ Participants are presented with two different hypothetical scenarios, known as a choice set, which vary in the levels of the attributes, and are asked to indicate their preference (scenario a, scenario b, both the same). Researchers can then identify participant preferences for the levels of each attribute and the relative weight of each attribute in participant preferences. This approach is increasingly used in health policy research.

### Measures

#### Discrete choice experiment

##### Attribute selection

Attributes for investigation were based on a recent systematic review conducted by our team,^13^ and on those which are likely to be considered by policymakers if a mandatory vaccination scheme were to be proposed (see supplementary materials for detailed rationale). For this study, we selected six attributes, four attributes with two levels each (vaccine [6-in-1, MMR], child age group [2 years and older, 5 years and older], ability to opt out [medical exemptions only, medical and religious belief exemptions], availability of a compensation scheme [offered, not offered]) and two attributes with three levels each (incentive [no incentive, £130 cash pay-out for parent per vaccine dose, £130 voucher for child per vaccine dose], penalty [£450 fine for each dose of the vaccine missed; child unable to attend school or childcare if unvaccinated, parent not able to claim Child Benefits^1^ if child not vaccinated]; Table 1).

**Table 1.**
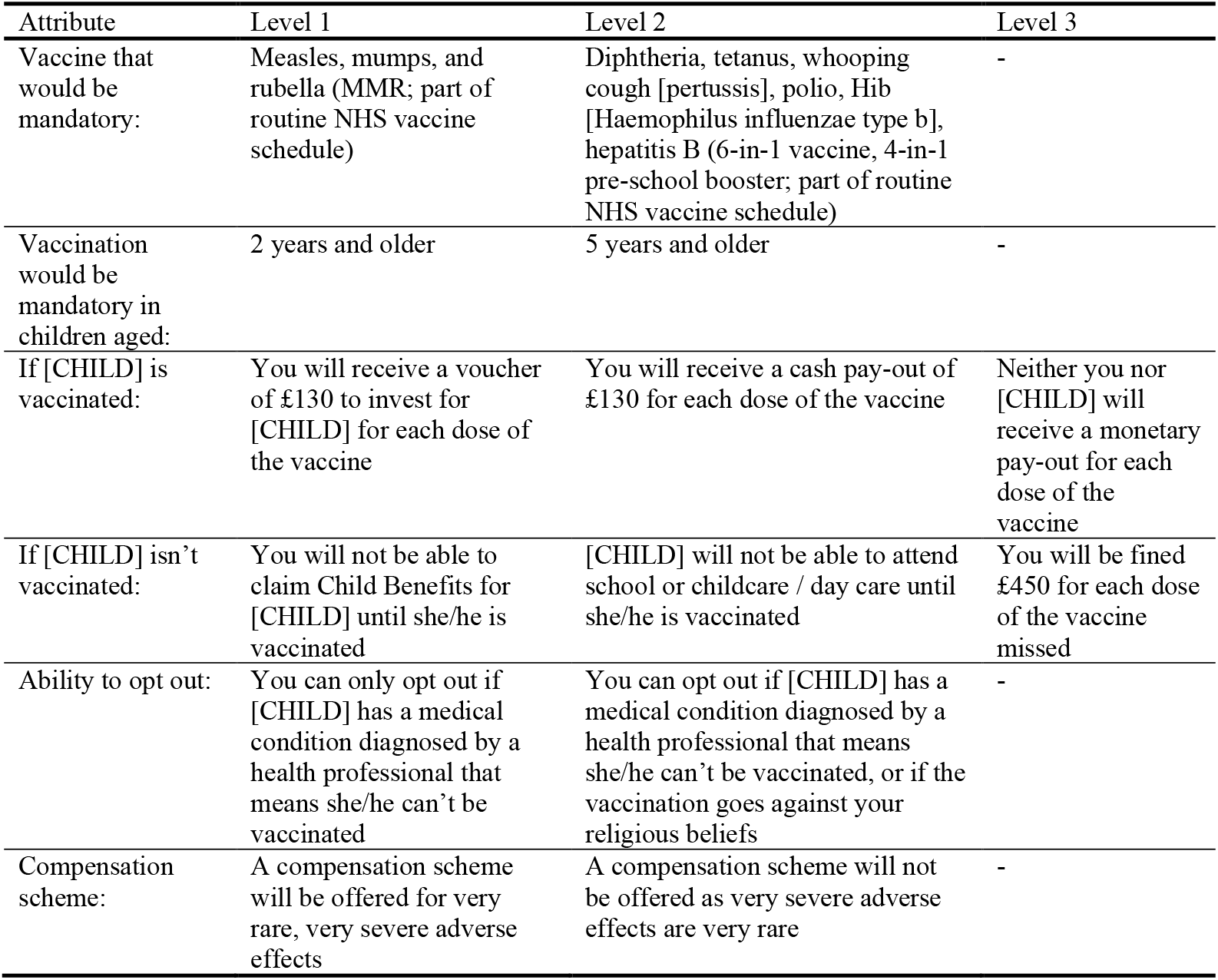
Attributes and levels included in the discrete choice experiment.

##### Experimental design

For this study, a full factorial design would require participants to rate every scenario (3^2^x2^4^=144); this was not practical. We used a fractional factorial design with each participant rating sixteen choice sets. The sample was split, with half receiving group set A and half receiving group set B. Thus, we used 32 choice sets (total of 64 scenarios). The design was optimised using a modified Fedorov algorithm (D-optimisation) to determine the 64 most optimal allocations, and 32 most optimal comparisons from the complete factorial design.^14,15^ Within each group set, choice sets were randomly presented to avoid order effects.

For each choice set, participants were asked which of the scenarios for a hypothetical mandatory vaccination scheme they preferred (scenario 1 or 2). Participants could also indicate that they did not prefer either scenario by choosing a third opt-out option (“neither”). Where participants selected that they did not prefer either scenario, they were subsequently asked to indicate which scenario was most preferable (“the least bad”). Figure 1 shows an example choice set.

**Figure 1.**
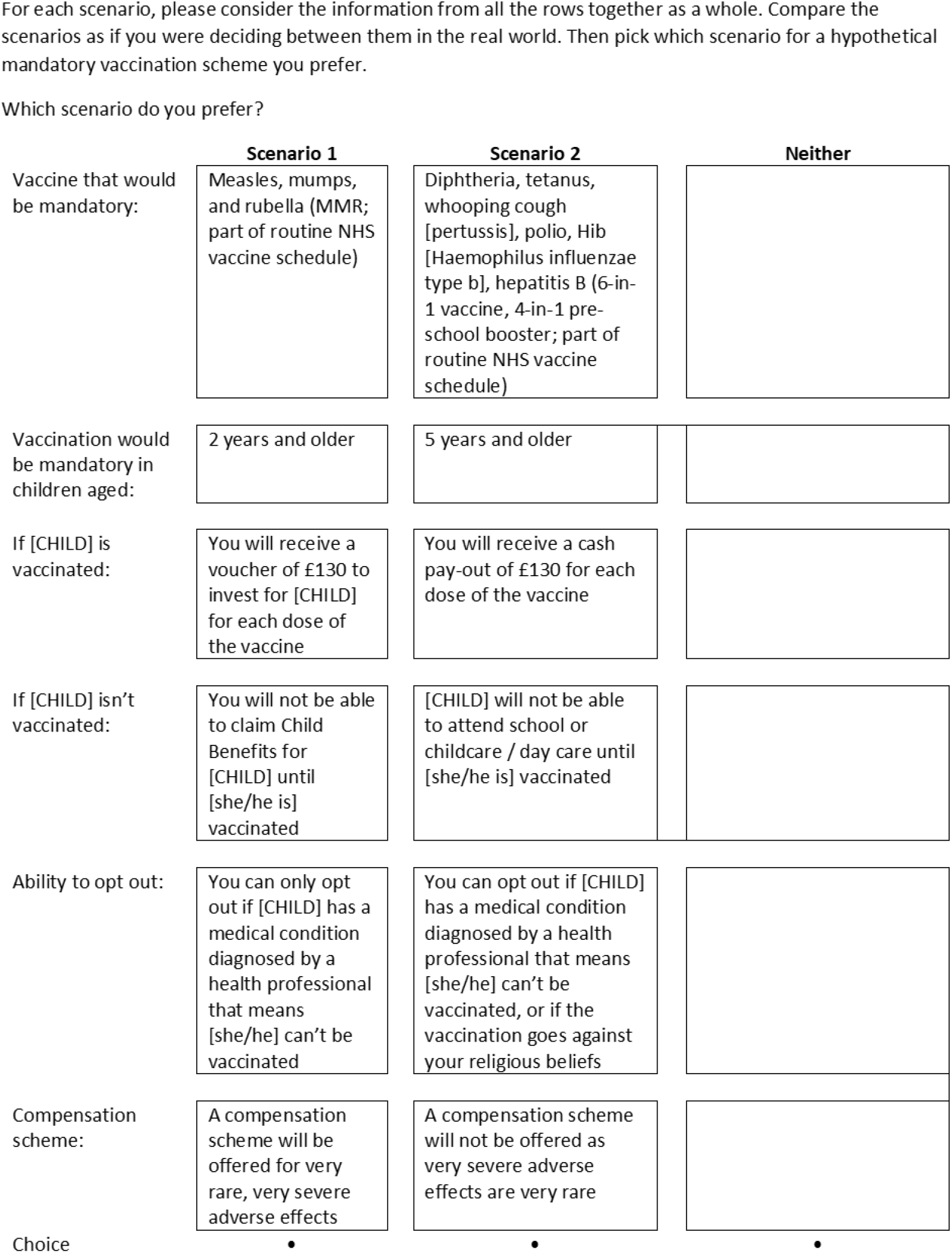
Example choice set

Internal validity was measured using a repeated question to check the stability of responses.^16^ We randomly selected a choice set from group set A and group set B, which was presented to participants at the end of the group set. The choice set was inverted (scenario 1 became scenario 2, and vice versa). Thus, participants rated a total of seventeen choice sets. Results from the internal validity choice set were not included in DCE analyses.

Final survey materials are presented in the supplementary materials.

#### Psychological factors

Participants were asked to what extent they agreed with a series of eleven statements on an eleven-point Likert scale (“strongly disagree (0)” to “strongly agree (10)”). Statements measured theoretically driven constructs associated with uptake of vaccines and perceptions about mandatory vaccination including perceived susceptibility and severity of vaccine preventable illnesses, perceived effectiveness and safety of child vaccines, approval of mandatory vaccination in general, thinking that vaccination campaigns are about making money for the manufacturers, preference for natural immunity and herd immunity. Statements were adapted from existing literature.^10,17,18^

#### Socio-demographic characteristics

We asked participants for their age, sex, ethnicity, highest educational qualification, employment status, total household income, marital status, how many children they were the parent or legal guardian of, and whether they had a chronic illness.

For the questionnaire, we asked participants to think about one of their children who was aged 5 years or younger (index child). We collected data about this child’s age (in years), sex, and whether they had a chronic illness.

Participants provided their full postcode, from which region and index of multiple deprivation (2019) were derived.

### Pilot testing

We piloted the survey, including the DCE, with five parents of children aged 5 years or younger (three female, two male) for understanding of materials. Comments from pilot participants resulted in framing the DCE with reference to a named index child, changes in wording to the study materials and changes in the layout of the DCE.

### Data collection

#### Design

Data were collected using an online cross-sectional survey (conducted 20 May to 7 June 2021) conducted by Panelbase, a Market Research Society Company Partner.

#### Participants

Participants were recruited from two specialist research panel providers (people who have opted in to take part in online surveys; Panelbase and Lucid). Those aged 18 years or older, living in England, with a child aged 5 years or younger were eligible. Recruitment used non-probability sampling, an approach common in standard opinion polling methods. Quota sampling was based on sex, ethnicity and Government Office Region to ensure the sample was broadly representative of the English general population.

#### Power

A full sample size calculation would require estimates of the true values of the parameters, which are not known before beginning the research. Estimates indicate that a reliable model can be achieved with a minimum of twenty participants per choice set.^19^ Given 32 choice sets, we aimed to recruit 1000 participants (32×30=960), to give us adequate power for model convergence in the main analysis.

### Ethics

Ethical approval for this study was granted by King’s College London Psychiatry, Nursing, and Midwifery Research Ethics subcommittee (reference number LRS-20/21-21880).

### Data Analysis

Socio-demographic characteristics of participants were analysed by index child vaccination status (not, partially or fully vaccinated).

To check D-efficient design properties of the DCE, sample attribute level balance summary statistics were calculated, orthogonality of the design was assessed by inspection of the correlation matrix, and choice sets were observed visually to inspect the overlap.

#### DCE Analysis (Mixed-effect logit regression)

Mixed effects conditional logit regression models were run to investigate parental preferences for each level of individual attributes, fitting the attributes as random effects, with 500 Halton draws.^12,20^ We calculated conditional preference for levels within the attributes and comparisons of relative importance of attributes, log likelihood, likelihood ratio test and the Akaike information criterion (AIC) for goodness of fit of the model. These analyses were run in Stata version 16.

#### Subgroup Analyses

We identified groups in whom uptake of child vaccination and approval of mandatory vaccination were lower based on parent socio-demographic characteristics. Groups and cut-offs were identified *a-priori* based on the literature (lower parental education [other or no qualifications, vs degree or higher], lower total household income [up to £29,999, vs £30,000 or higher], greater number of children [one, two, three or more; proxy for household size], not identifying as white ethnicity [white, vs other], living in London [London, vs the North (North East, North West, Yorkshire and the Humber), the midlands (East Midlands, West Midlands, East of England), the South (South East, South West)], not being partnered [not partnered, vs partnered], younger parent age [under 30 years, vs 30 years and over]).^3,9,13,21-25^ We also grouped parents by index of multiple deprivation using a three-way split (lowest [living in most deprived deciles, 1 to 3], vs middle [living in deciles, 4 to 7], highest [living in least deprived deciles, 8 to 10].

We used dimension reduction techniques to aid feature identification for psychological factors. We conducted an exploratory factor analysis, using direct oblimin rotation as items may have been correlated. All psychological factor items were included in the factor analyses. The number of factors was determined using a scree plot. We created a single variable (“approval of child vaccination”) by summing variables that loaded strongly onto the first component (range 0 to 60), with higher scores reflecting more positive vaccination beliefs. We hypothesised that people with more positive vaccine sentiments would be more likely to vaccinate their child and to approve of mandatory vaccination schemes. To group participants *a-priori*, we used a tertile split (approval of child vaccination: lowest [scores 0 to 42], middle [scores 43 to 52], highest [scores 53 to 60]). SPSS version 26 was used for these analyses.

DCE analyses were repeated within each subgroup.

## Results

### Participants

1,056 participants completed the survey, but 55 were excluded for quality assurance purposes (e.g. completing the survey too quickly, or giving the same answer to several consecutive questions). Therefore, the final sample was 1,001. Participants were broadly representative of the English population (53% female, 84% white ethnicity) and ranged in age between 18 and 65 years (mean=33·6 years, SD=7·1). Participant characteristics differed by child vaccination status, with female, older, white, and partnered parents being more likely to report that their child was fully vaccinated (Table 2). People who lived in London were less likely to have a fully vaccinated child, as were those whose child had a chronic illness.

**Table 2.**
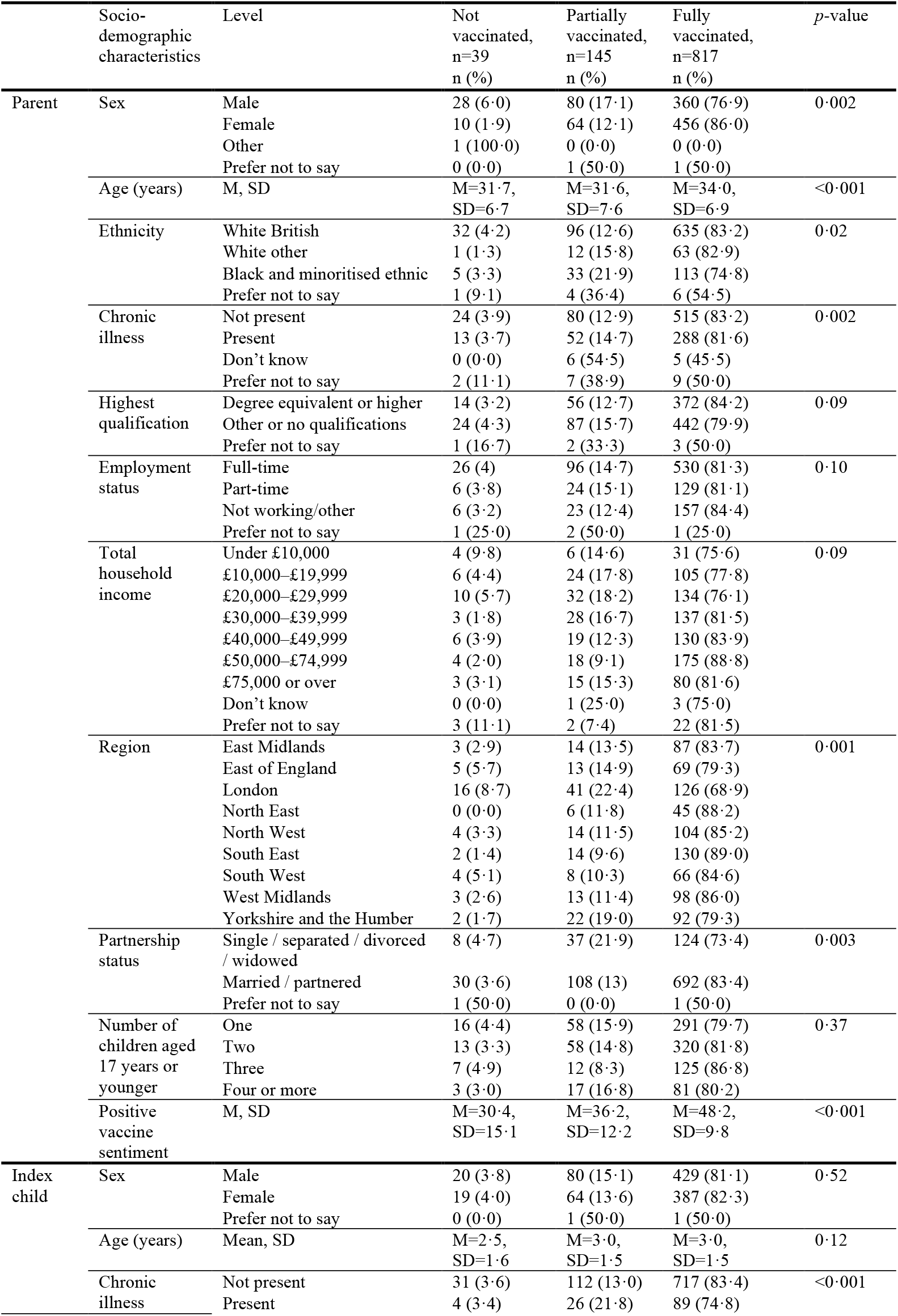
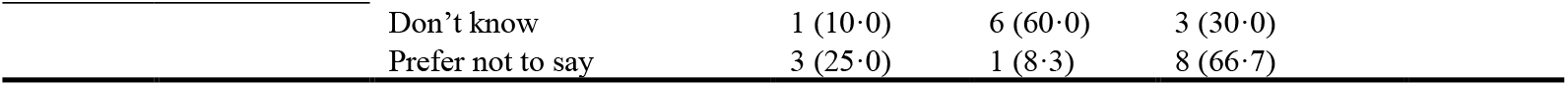
Participant characteristics, by index child vaccination status

|  | Socio-demographic characteristics | Level | Not vaccinated, n=39<br>n (%) | Partially vaccinated, n=145<br>n (%) | Fully vaccinated, n=817<br>n (%) | p-value |
| --- | --- | --- | --- | --- | --- | --- |
| Parent | Sex | Male | 28 (6.0) | 80 (17.1) | 360 (76.9) | 0.002 |
|  |  | Female | 10 (1.9) | 64 (12.1) | 456 (86.0) |  |
|  |  | Other | 1 (100.0) | 0 (0.0) | 0 (0.0) |  |
|  |  | Prefer not to say | 0 (0.0) | 1 (50.0) | 1 (50.0) |  |
|  | Age (years) | M, SD | M=31.7,<br>SD=6.7 | M=31.6,<br>SD=7.6 | M=34.0,<br>SD=6.9 | <0.001 |
|  | Ethnicity | White British | 32 (4.2) | 96 (12.6) | 635 (83.2) | 0.02 |
|  |  | White other | 1 (1.3) | 12 (15.8) | 63 (82.9) |  |
|  |  | Black and minoritised ethnic | 5 (3.3) | 33 (21.9) | 113 (74.8) |  |
|  |  | Prefer not to say | 1 (9.1) | 4 (36.4) | 6 (54.5) |  |
|  | Chronic illness | Not present | 24 (3.9) | 80 (12.9) | 515 (83.2) | 0.002 |
|  |  | Present | 13 (3.7) | 52 (14.7) | 288 (81.6) |  |
|  |  | Don't know | 0 (0.0) | 6 (54.5) | 5 (45.5) |  |
|  |  | Prefer not to say | 2 (11.1) | 7 (38.9) | 9 (50.0) |  |
|  | Highest qualification | Degree equivalent or higher | 14 (3.2) | 56 (12.7) | 372 (84.2) | 0.09 |
|  |  | Other or no qualifications | 24 (4.3) | 87 (15.7) | 442 (79.9) |  |
|  |  | Prefer not to say | 1 (16.7) | 2 (33.3) | 3 (50.0) |  |
|  | Employment status | Full-time | 26 (4) | 96 (14.7) | 530 (81.3) | 0.10 |
|  |  | Part-time | 6 (3.8) | 24 (15.1) | 129 (81.1) |  |
|  |  | Not working/other | 6 (3.2) | 23 (12.4) | 157 (84.4) |  |
|  |  | Prefer not to say | 1 (25.0) | 2 (50.0) | 1 (25.0) |  |
|  | Total household income | Under £10,000 | 4 (9.8) | 6 (14.6) | 31 (75.6) | 0.09 |
|  |  | £10,000–£19,999 | 6 (4.4) | 24 (17.8) | 105 (77.8) |  |
|  |  | £20,000–£29,999 | 10 (5.7) | 32 (18.2) | 134 (76.1) |  |
|  |  | £30,000–£39,999 | 3 (1.8) | 28 (16.7) | 137 (81.5) |  |
|  |  | £40,000–£49,999 | 6 (3.9) | 19 (12.3) | 130 (83.9) |  |
|  |  | £50,000–£74,999 | 4 (2.0) | 18 (9.1) | 175 (88.8) |  |
|  |  | £75,000 or over | 3 (3.1) | 15 (15.3) | 80 (81.6) |  |
|  |  | Don't know | 0 (0.0) | 1 (25.0) | 3 (75.0) |  |
|  |  | Prefer not to say | 3 (11.1) | 2 (7.4) | 22 (81.5) |  |
|  | Region | East Midlands | 3 (2.9) | 14 (13.5) | 87 (83.7) | 0.001 |
|  |  | East of England | 5 (5.7) | 13 (14.9) | 69 (79.3) |  |
|  |  | London | 16 (8.7) | 41 (22.4) | 126 (68.9) |  |
|  |  | North East | 0 (0.0) | 6 (11.8) | 45 (88.2) |  |
|  |  | North West | 4 (3.3) | 14 (11.5) | 104 (85.2) |  |
|  |  | South East | 2 (1.4) | 14 (9.6) | 130 (89.0) |  |
|  |  | South West | 4 (5.1) | 8 (10.3) | 66 (84.6) |  |
|  |  | West Midlands | 3 (2.6) | 13 (11.4) | 98 (86.0) |  |
|  |  | Yorkshire and the Humber | 2 (1.7) | 22 (19.0) | 92 (79.3) |  |
|  | Partnership status | Single / separated / divorced / widowed | 8 (4.7) | 37 (21.9) | 124 (73.4) | 0.003 |
|  |  | Married / partnered | 30 (3.6) | 108 (13) | 692 (83.4) |  |
|  |  | Prefer not to say | 1 (50.0) | 0 (0.0) | 1 (50.0) |  |
|  | Number of children aged 17 years or younger | One | 16 (4.4) | 58 (15.9) | 291 (79.7) | 0.37 |
|  |  | Two | 13 (3.3) | 58 (14.8) | 320 (81.8) |  |
|  |  | Three | 7 (4.9) | 12 (8.3) | 125 (86.8) |  |
|  |  | Four or more | 3 (3.0) | 17 (16.8) | 81 (80.2) |  |
|  | Positive vaccine sentiment | M, SD | M=30.4,<br>SD=15.1 | M=36.2,<br>SD=12.2 | M=48.2,<br>SD=9.8 | <0.001 |
| Index child | Sex | Male | 20 (3.8) | 80 (15.1) | 429 (81.1) | 0.52 |
|  |  | Female | 19 (4.0) | 64 (13.6) | 387 (82.3) |  |
|  |  | Prefer not to say | 0 (0.0) | 1 (50.0) | 1 (50.0) |  |
|  | Age (years) | Mean, SD | M=2.5,<br>SD=1.6 | M=3.0,<br>SD=1.5 | M=3.0,<br>SD=1.5 | 0.12 |
|  | Chronic illness | Not present | 31 (3.6) | 112 (13.0) | 717 (83.4) | <0.001 |
|  |  | Present | 4 (3.4) | 26 (21.8) | 89 (74.8) |  |

|  |  |  |  |
| --- | --- | --- | --- |
| Don't know | 1 (10·0) | 6 (60·0) | 3 (30·0) |
| Prefer not to say | 3 (25·0) | 1 (8·3) | 8 (66·7) |

### DCE

The attributes that most influenced participants’ preferences were incentives and penalties included in mandatory vaccination schemes, followed by the availability of a compensation scheme (Table 3). Participants had a strong preference for mandatory vaccination schemes that offered a financial incentive to themselves or their child for each vaccine dose, compared to not receiving an incentive. When using incentivisation to the child as the reference category, participants slightly preferred a cash incentive for themselves, compared to an incentive to their child. Compared to receiving a £450 fine for each dose of the vaccine missed, parents preferred mandatory vaccination schemes that did not allow unvaccinated children to attend school or day care and those that withheld financial benefits for parents of unvaccinated children. When denying unvaccinated children schooling or childcare was denoted as the reference category, there was no significant preference for schemes that withheld benefits for parents of unvaccinated children. Participants preferred mandatory vaccination schemes that: offered a compensation scheme, versus not offering one; mandated the 6-in-1, rather than MMR vaccine; mandated vaccination in children aged 2 years and older, compared to 5 years and older; and had a slight preference for schemes with medical exemptions only, rather than those with medical and religious exemptions.

**Table 3.** Percentage of participants who have a positive preference for a mandatory vaccine scheme attribute.

| Attribute | Level | Mean<br>(standard<br>error) | % who prefer this level (95%<br>CI) | p-level | Relative<br>importance |
| --- | --- | --- | --- | --- | --- |
| Vaccine | MMR<br>6-in-1 | 0.13 (0.03) | Reference<br>58.2 (54.6 to 61.7) | <0.001 | 10.5% |
| Child age group | 2 years and older<br>5 years and older | -0.14 (0.03) | Reference<br>42.6 (39.4 to 45.9) | <0.001 | 11.4% |
| Incentive | No incentive<br>Incentive for child (£130 voucher)<br>Incentive for parent (£130 cash pay-out) | 0.32 (0.03)<br>0.39 (0.03) | Reference<br>74.8 (71.0 to 78.3)<br>80.9 (76.3 to 85.0) | <0.001<br><0.001 | 30.7% |
|  | Incentive for child (£130 voucher)<br>Incentive for parent (£130 cash pay-out) | 0.06 (0.03) | Reference<br>56.3 (50.0 to 62.4) | 0.05 |  |
| Penalty | £450 fine<br>Parent not able to claim Child Benefits for child until they are<br>vaccinated<br>Child not able to attend school or childcare / day care until they<br>are vaccinated | 0.29 (0.03)<br>0.32 (0.03) | Reference<br>67.6 (63.6 to 71.4)<br>69.5 (65.7 to 73.1) | <0.001<br><0.001 | 25.4% |
|  | Child not able to attend school / day care until they are vaccinated<br>Parent not able to claim Child Benefits for child until they are<br>vaccinated | -0.04 (0.03) | Reference<br>47.5 (42.8 to 52.2) | 0.30 |  |
| Ability to opt out | Medical exemption<br>Medical and religious belief exemption | -0.05 (0.03) | Reference<br>45.5 (41.1 to 50.0) | 0.05 | 4.0% |
| Compensation<br>scheme | Not offered<br>Offered | 0.23 (0.03) | Reference<br>66.4 (62.7 to 69.8) | <0.001 | 18.1% |

Model fit summary statistics. Log likelihood = -10,313.87, Likelihood Ratio Test = 1,030.58, $p < 0.0001$ , Akaike's Information Criterion = 20,661.73

In group set A, internal consistency was 63% (n=298/473), whereas in group set B, internal consistency was 67% (n=355/528).

### Subgroup Analyses

Subgroup analyses followed the same pattern as in the whole sample. Major differences in subgroups are described narratively; all results are reported in the supplementary materials.

#### Socio-demographic characteristics

Participants from black and minoritized ethnic groups showed different patterns of preferences to white participants, placing more importance on the ability to opt out of a mandatory vaccination scheme (still preferred schemes only allowed medical exemptions) and that offered a compensation scheme; less emphasis was placed on which vaccine and age group the vaccine would be made mandatory (no preference).

There were some regional differences in preferences for mandatory vaccination schemes. Participants living in the South placed less emphasis on incentives and penalties of vaccination schemes but placed more importance on the ability to opt out (schemes allowing only medical exemptions preferred). Participants living in London also gave more importance on the ability to opt out (medical exemptions only preferred), and less importance to the vaccine to be made mandatory (no preference).

Participants who reported not having a partner also placed more emphasis on the ability to opt out of a mandatory vaccination scheme; these participants preferred schemes that offered medical and religious belief exemptions. Less emphasis was placed on the vaccine to be made mandatory (no preference).

Participants with three or more children placed a much stronger emphasis on incentives for vaccination, and less emphasis on the age group in which vaccine would be mandated (no preference). Parents aged under 30 did not show a preference for schemes that would not allow them to claim benefits for their child if not vaccinated.

#### Psychological factors

Dimension reduction techniques indicated that psychological factors could be grouped into two distinct groups, with the first component being the most influential (supplementary materials). The first component reflected generally positive vaccine sentiments (“approval of child vaccines”; child vaccines are effective and safe, approval of mandatory vaccination), while the second reflected generally negative vaccine sentiments (“disapproval of child vaccines”; child vaccines cause side effects, not liking child vaccines generally, preferring natural immunity, and thinking that child vaccination campaigns are financially motivated). Factor loadings of individual items onto components are reported in the supplementary materials.

Preferences for a mandatory vaccination scheme varied by vaccine sentiment. Those with the most positive vaccine sentiments placed less emphasis on incentives and penalties of the schemes, and more emphasis on the vaccine to be made mandatory and the age group in which vaccines should be made mandatory; there was no difference in direction of preference in this group. Participants with the least positive vaccine sentiments showed a preference for mandatory vaccination schemes that allowed medical and religious belief exemptions and those that mandated vaccination in older children.

## Discussion

We investigated English parents’ preferences for a mandatory vaccination scheme. The attributes that most strongly influenced parental preferences were financial incentives for vaccination, the penalty imposed for missing a vaccine dose, and the inclusion of a compensation scheme for severe adverse effects. Our results contrast with findings that parents dislike mandatory vaccination schemes that offer financial incentives.^13^ This difference is likely due to selective sampling used in qualitative studies that were synthesised. A previous DCE in English parents found a preference for financial incentives only in parents “at high risk” of vaccine refusal (living in more deprivation, having a child with a chronic illness, being a single parent, being aged less than 20 years, and having three or more children).^10,11^ However, this study included fewer parents in each subgroup and so had less power to detect smaller differences.

We found that parents preferred schemes that denied access to schooling and childcare, or that stopped parents of unvaccinated children receiving tax benefits, compared to receiving a fine for each vaccine dose mixed. Systematic review findings indicating that parents felt peace of mind in schemes that restricted mixing of unvaccinated children at school or day care^13^ suggest that, should a mandatory vaccination scheme be proposed, this may be the preferential option. However, caution should be taken as parents who strongly oppose vaccination may seek alternative ways to school their child such as home-schooling.^26^ Care should also be taken to avoid penalizing the child for their parents’ vaccination decision.^13^

Implementing a mandatory vaccination scheme is unlikely to affect vaccination decisions in parents of children who vaccinate their child voluntarily. Instead, their aim is to increase uptake in those who refuse vaccines for their children. We investigated parental preferences in subgroups that were less likely to vaccinate their child and have less positive attitudes towards mandatory vaccination. There were few meaningful differences between subgroup and whole sample analyses. However, parents with the least positive vaccine beliefs using our composite measure displayed significant differences in their preferences, preferring schemes that allowed medical and religious belief exemptions (versus medical exemptions only) and that mandated vaccination in older (versus younger) children. Previous research indicates that mandating vaccination has the potential to increase vaccine refusal in children of vaccine hesitant parents,^27^ and entrench negative vaccine beliefs.^26^ It is possible that implementing a mandatory vaccination scheme that does not align with parents’ preferences may lead to more negative vaccine beliefs and refusal. Therefore, the decision to implement a mandatory vaccine scheme should be taken with caution.

Parent-reported child vaccine uptake in this study broadly followed patterns seen in other literature and official national statistics, with lower uptake of vaccines in children of younger parents, single parents, parents of black and minoritized ethnic groups, and parents living in London.^3,21,23^ Men also reported lower uptake of vaccination in their children; we are unsure why this may be. Children with a chronic illness were also less likely to be vaccinated. This may be due to contraindications for vaccination. Parents with less positive vaccine sentiments also reported lower child vaccine uptake. Parents may not vaccinate their child due to lack of access to vaccination or not wanting to vaccinate.^28^ While mandating vaccination may affect parents’ willingness to vaccinate their child, they are unlikely to address access problems. If the decision to implement a mandatory vaccination scheme is made, care should be taken to ensure that it does not disproportionately affect disadvantaged groups.^6^

Strengths of this study include the use of a large sample of parents, whose socio-demographic characteristics (sex, ethnicity, region) were broadly reflective of the general English population. As quota sampling uses targets based on socio-demographic characteristics and prevents people from completing the survey if targets have already been fulfilled, response rate is not an accurate measurement of response bias. Limitations include that power to detect differences in subgroup analyses was lower than in whole-sample analyses. We are unsure whether the beliefs and attitudes of those who sign up to take part in online research are representative of the beliefs and attitudes of the general population. However associations within the data should remain valid.^29^ To minimise biasing of the sample towards those interested in vaccination, participants did not know the topic of the survey when choosing to take part. Internal consistency for the sample was 63% (group A) to 67% (group B). This is likely because scenarios were complex. Previous research suggests that excluding participants who do not answer consistently may bias the sample, as well as removing valid preferences from the data and reducing the power of the experiment.^30^ We presented participants with sixteen choice sets as part of the DCE, under the maximum recommended, to reduce the influence of research fatigue.^12^ Due to the COVID-19 pandemic and additional burden that public health officials and policy makers were under, we were unable to conduct qualitative interviews to inform our DCE materials. Results may not be generalisable to other countries or vaccines.

While mandatory vaccination schemes have been implemented in several countries, the cultural context is likely to determine their sucess.^7^ Results of this study should be used to inform policymakers’ decisions about how best to implement a mandatory vaccination scheme in England in the eventuality that one is proposed. Results suggest that direct financial rewards were key drivers to English parents’ preferences for a mandatory vaccination scheme, followed by the indirect financial security of having a compensation scheme in place for complications stemming from the vaccine. However, preferences differed by vaccine sentiment.

## Data Availability

Anonymised data, and an accompanying data dictionary will be made available to others after publication, beginning 3 months and ending 5 years following article publication. Researchers who provide a statistical analysis plan addressing a methodological robust legitimate research question will be able to request the data to achieve the aims in the approved proposal. Proposals should be directed to to gain access. Data requestors will need to sign a data access agreement.

## Acknowledgements

The authors would like to thank Professor G James Rubin for his help with the study.

## Author contributions

LS and BC conceptualised, acquired funding for the study, curated and validated data, as well as contributing to the study methodology. Both authors contributed to the original draft of the manuscript. LS is the guarantor of these findings.

## Footnotes

1 In England, parents can receive a Child Benefit allowance for children under 16 years old (https://www.gov.uk/child-benefit).

