## Supplementary materials for "Parental preferences for a mandatory vaccination scheme in England; a discrete choice experiment"

### Supplementary materials 1. Selection of vaccine attributes

The first attribute we decided to investigate was the choice of vaccine to be made mandatory (“vaccine”). We based levels on the two vaccines with the lowest uptake in England in recent years, namely the measles, mumps, and rubella (MMR) vaccine, and the 6-in-1 vaccine (diphtheria, tetanus, whooping cough [pertussis], polio, Hib [Haemophilus influenzae type b], hepatitis B), aside from the child influenza vaccine.[1] The child influenza vaccine was excluded because it is given annually, rather than as a set number of doses (MMR = 2, 6-in-1 vaccine = 3) and so other issues (e.g. regularity of vaccination) may affect uptake and influence comparability of other parameters (e.g. financial incentives).

We chose levels of our second attribute (“child age group”) based on groupings used by the NHS to measure vaccine uptake (2 years and 5 years).

The third attribute investigated was incentives for vaccination. One study investigating parental preferences for a quasi-mandatory vaccination scheme investigated the use of financial incentives for vaccination, finding that in parents who required a reward for vaccinating their child, the average minimum “willingness to accept” amount was £112.[2] We used an inflation calculator to account for inflation between 2014 (when the previous study was conducted) and 2020 (the most up-to-date estimates available at the time), to give an incentive amount of £130.[3] We compared financial incentives to the child (as a voucher to invest for the child), to the parent (as a cash pay-out; parents prefer a cash incentive to a voucher incentive[2,4]), and no financial incentive.

Penalties for not having vaccinated one’s child were our fourth attribute. Globally, educational penalties, where the child cannot attend school, child care or day care until vaccinated, and financial penalties, either through a monetary fine or not being able to claim benefits for one’s child if not vaccinated, are the most commonly used.[5] Therefore, we selected levels of our attribute to reflect an educational penalty, and two types of a financial penalty: not being able to claim Child Benefits and receiving a fine. Fines have been shown to increase vaccine uptake, with evidence suggesting that for each 500 euro increase in fine, there was an increase of 0.8% for measles vaccine coverage and 1.1% increase in pertussis vaccine coverage.[6] We converted this amount to Great British Pounds using the conversion rate available at the time to give a fine amount of £450.

Our fifth attribute was the ability to opt out of the mandatory vaccination scheme. Evidence, much of it from the United States, indicates that non-medical exemptions, such as exemptions based on religious or personal beliefs affect vaccine uptake.[7] We investigated the preferences for allowing exemptions based solely on medical grounds, or on medical and religious grounds.

The presence of a compensation scheme was our sixth attribute (offered, versus not offered). This was based on findings of parental safety concerns about not being offered a compensation scheme if vaccines were mandated.[8]

### Supplementary materials 2. Full survey materials

*Text in italics not shown to participants*

**Q1.** Which country do you live in? *[single code]*

- England
- Scotland
- Wales
- Northern Ireland
- Outside the UK

*Screen out if not England*

**Q2.** This survey requires respondents to give their full postcode. Are you happy to provide this? *[single code]*

This information will only be used for statistical purposes to analyse the results by specific areas, such as Local Authority, Constituency and Government areas. Asking for your postcode saves you time and helps us to report more accurate information. All answers will be treated entirely anonymously and postcode information will not be used for any other purpose.

- Yes
- No

*Screen out if No*

**Q3.** Could you please provide your full English postcode? Please ensure to include a space where applicable, e.g. AB1 2CD

- Free text insertion

**Q4.** What is your age (in years)?

- Insert number

*Screen out if aged 17 or under*

**Q5.** Do you identify as: *[single code]*

- Male
- Female
- Prefer to self-describe
- Prefer not to say

**Q6.** Which of the following best describes you? *[single code]*

White

- British
- Irish
- Eastern European
- Any other white background

Mixed

- White and Black Caribbean
- White and Black African
- White and Asian
- Any other mixed background

Asian or Asian British

- Indian
- Pakistani
- Bangladeshi

- Sri Lankan
- Any other Asian background

Black or Black British

- Caribbean
- African
- Any other Black background

Chinese or Other ethnic group

- Chinese
- Any other
- Prefer not to say

**Q7.** How many children, if any, of the following ages, are you the parent or legal guardian of? (Please select one option on each row) *[single code]*

| | 0 | 1 | 2 | 3 | 4 | 5 | 6 or more |
|---|---|---|---|---|---|---|---|
| 0 to 5 years old | | | | | | | |
| 6 to 17 years old | | | | | | | |

*Screen out if 0 to 5 years =0*

**Q8.** For this questionnaire, we would like you to think about **one of your children who is aged 5 years or younger**. If you have more than one child who is aged 5 years or younger, please answer about your child with the most recent birthday.

Can you tell us the first name of your child. This is just so we can refer to them throughout the survey. You can give a fake name for them if you want:

Free-text insertion

**Q9.** How old is [CHILD] (in years)? *[single code]*

- 0
- 1
- 2
- 3
- 4
- 5
- Older than 5 years

*Screen out if "older than 5 years"*

**Q10.** Is [CHILD] a: *[single code]*

- Girl
- Boy
- Prefer not to say

The following questions are going to be about child vaccinations. In England, parents can choose whether to vaccinate their child. In some countries, child vaccination is mandatory. Mandatory vaccination laws across the world differ widely.

In the next questions, we are going to show you two hypothetical scenarios for a mandatory vaccination scheme. We would like you to tell us which scenario you prefer.

For each scenario, please consider the information from all the rows together as a whole. Compare the scenarios as if you were deciding between them in the real world. Then pick which scenario for a hypothetical mandatory vaccination scheme you prefer.

You can state that you do not prefer either scenario by choosing 'Neither'. If you choose 'Neither' as your preferred option then we would still like you to indicate whether scenario 1 or 2 would be the most preferable to you *(i.e., the least bad)*.

An example is shown below:

Which scenario do you prefer?

| | **Scenario 1** | **Scenario 2** | **Neither** |
|---|---|---|---|
| Vaccine that would be mandatory: | Measles, mumps, and rubella (MMR; part of routine NHS vaccine schedule) | Diphtheria, tetanus, whooping cough [pertussis], polio, Hib [Haemophilus influenzae type b], hepatitis B (6-in-1 vaccine, 4-in-1 pre-school booster; part of routine NHS vaccine schedule) | |
| Vaccination would be mandatory in children aged: | 2 years and older | 5 years and older | |
| If [CHILD] is vaccinated: | You will receive a voucher of £130 to invest for [CHILD] for each dose of the vaccine | You will receive a cash pay-out of £130 for each dose of the vaccine | |
| If [CHILD] isn't vaccinated: | You will not be able to claim Child Benefits for [CHILD] until [she/he is] vaccinated | [CHILD] will not be able to attend school or childcare / day care until [she/he is] vaccinated | |
| Ability to opt out: | You can only opt out if [CHILD] has a medical condition diagnosed by a health professional that means [she/he] can't be vaccinated | You can opt out if [CHILD] has a medical condition diagnosed by a health professional that means [she/he] can't be vaccinated, or if the vaccination goes against your religious beliefs | |
| Compensation scheme: | A compensation scheme will be offered for very rare, very severe adverse effects | A compensation scheme will not be offered as very severe adverse effects are very rare | |
| **DCE Example.** Choice *[single code]* | ◦ | ◦ | ◦ |

*If Neither selected display:*

We would still like you to indicate whether Scenario 1 or 2 is most preferable to you (i.e., the least bad) – which would you choose?

| | **Scenario 1** | **Scenario 2** |
|---|---|---|
| Vaccine that would be mandatory: | Measles, mumps, and rubella (MMR; part of routine NHS vaccine schedule) | Diphtheria, tetanus, whooping cough [pertussis], polio, Hib [Haemophilus influenzae type b], hepatitis B (6-in-1 vaccine, 4-in-1 pre-school booster; part of routine NHS vaccine schedule) |
| Vaccination would be mandatory in children aged: | 2 years and older | 5 years and older |
| If [CHILD] is vaccinated: | You will receive a voucher of £130 to invest for [CHILD] for each dose of the vaccine | You will receive a cash pay-out of £130 for each dose of the vaccine |
| If [CHILD] isn't vaccinated: | You will not be able to claim Child Benefits for [CHILD] until [she/he is] vaccinated | [CHILD] will not be able to attend school or childcare / day care until [she/he is] vaccinated |
| Ability to opt out: | You can only opt out if [CHILD] has a medical condition diagnosed by a health professional that means [she/he] can't be vaccinated | You can opt out if [CHILD] has a medical condition diagnosed by a health professional that means [she/he] can't be vaccinated, or if the vaccination goes against your religious beliefs |
| Compensation scheme: | A compensation scheme will be offered for very rare, very severe adverse effects | A compensation scheme will not be offered as very severe adverse effects are very rare |
| **DCE Example a.** Choice *[single code]* | ◦ | ◦ |

Thank you for completing the example question. We will now show you a series of questions.

*SPLIT SAMPLE – half to Block A (Q1A to Q16A + IntConsistA), half to Block B (Q1B to Q16B + IntConsistB).*

*Block A. Order of DCE1A to DCE16A randomised (Internal Consistency check A anchored to end).*[1]

**DCE1A.** For each scenario, please consider the information from all the rows together as a whole. Compare the scenarios as if you were deciding between them in the real world. Then pick which scenario for a hypothetical mandatory vaccination scheme you prefer.

| Question number | Scenario | Vaccine | Child age group | Incentive | Penalty | Ability to opt out (exemptions) | Availability of compensation scheme |
|---|---|---|---|---|---|---|---|
| DCE1A | 1 | 6-in-1 | 5 years and older | No incentive | £450 fine | Medical exemptions only | Compensation scheme |
| | 2 | MMR | 2 years and older | Parent incentive | Restrictions on childcare | Medical and religious beliefs | No compensation scheme |
| DCE2A | 1 | MMR | 5 years and older | Child incentive | Restrictions on childcare | Medical and religious beliefs | Compensation scheme |
| | 2 | 6-in-1 | 2 years and older | No incentive | Restrictions on Child Benefits | Medical exemptions only | No compensation scheme |
| DCE3A | 1 | 6-in-1 | 2 years and older | No incentive | Restrictions on childcare | Medical and religious beliefs | Compensation scheme |
| | 2 | MMR | 5 years and older | Child incentive | Restrictions on Child Benefits | Medical exemptions only | No compensation scheme |
| DCE4A | 1 | 6-in-1 | 5 years and older | Child incentive | Restrictions on Child Benefits | Medical exemptions only | Compensation scheme |
| | 2 | MMR | 2 years and older | Parent incentive | £450 fine | Medical and religious beliefs | No compensation scheme |
| DCE5A | 1 | 6-in-1 | 2 years and older | No incentive | £450 fine | Medical exemptions only | Compensation scheme |
| | 2 | MMR | 5 years and older | Parent incentive | Restrictions on Child Benefits | Medical and religious beliefs | No compensation scheme |
| DCE6A | 1 | MMR | 2 years and older | Child incentive | Restrictions on childcare | Medical and religious beliefs | No compensation scheme |
| | 2 | 6-in-1 | 5 years and older | Parent incentive | £450 fine | Medical exemptions only | Compensation scheme |
| DCE7A | 1 | 6-in-1 | 5 years and older | No incentive | Restrictions on Child Benefits | Medical exemptions only | No compensation scheme |
| | 2 | MMR | 2 years and older | Child incentive | Restrictions on childcare | Medical and religious beliefs | Compensation scheme |
| DCE8A | 1 | 6-in-1 | 2 years and older | Child incentive | £450 fine | Medical exemptions only | Compensation scheme |
| | 2 | MMR | 5 years and older | No incentive | Restrictions on Child Benefits | Medical and religious beliefs | No compensation scheme |
| DCE9A | 1 | MMR | 2 years and older | No incentive | Restrictions on childcare | Medical exemptions only | Compensation scheme |
| | 2 | 6-in-1 | 5 years and older | Child incentive | £450 fine | Medical and religious beliefs | No compensation scheme |
| DCE10A | 1 | 6-in-1 | 2 years and older | Parent incentive | Restrictions on childcare | Medical exemptions only | No compensation scheme |
| | 2 | MMR | 5 years and older | Child incentive | Restrictions on Child Benefits | Medical and religious beliefs | Compensation scheme |
| DCE11A | 1 | 6-in-1 | 5 years and older | Child incentive | Restrictions on childcare | Medical exemptions only | No compensation scheme |

[1] DCE items displayed to participants as in example question above. For readability we have not included all items as shown to participants. Instead, we have included the combinations of attributes and levels shown to participants in each question. For exact wording of attributes and levels, see Table 1 of manuscript.

| | | | | | | | |
|---|---|---|---|---|---|---|---|
| | 2 | MMR | 2 years and older | No incentive | Restrictions on Child Benefits | Medical and religious beliefs | Compensation scheme |
| | 1 | MMR | 2 years and older | Child incentive | £450 fine | Medical exemptions only | No compensation scheme |
| DCE12A* | 2 | 6-in-1 | 5 years and older | Parent incentive | Restrictions on childcare | Medical and religious beliefs | Compensation scheme |
| DCE13A | 1 | 6-in-1 | 2 years and older | Parent incentive | Restrictions on Child Benefits | Medical exemptions only | No compensation scheme |
| | 2 | MMR | 5 years and older | No incentive | £450 fine | Medical and religious beliefs | Compensation scheme |
| DCE14A | 1 | MMR | 2 years and older | No incentive | £450 fine | Medical and religious beliefs | No compensation scheme |
| | 2 | 6-in-1 | 5 years and older | Child incentive | Restrictions on Child Benefits | Medical exemptions only | Compensation scheme |
| DCE15A | 1 | 6-in-1 | 2 years and older | Parent incentive | Restrictions on Child Benefits | Medical and religious beliefs | Compensation scheme |
| | 2 | MMR | 5 years and older | Child incentive | Restrictions on childcare | Medical exemptions only | No compensation scheme |
| DCE16A | 1 | 6-in-1 | 2 years and older | Child incentive | £450 fine | Medical exemptions only | No compensation scheme |
| | 2 | MMR | 5 years and older | Parent incentive | Restrictions on childcare | Medical and religious beliefs | Compensation scheme |
| Internal consistency check A | 1 | 6-in-1 | 5 years and older | Parent incentive | Restrictions on childcare | Medical and religious beliefs | Compensation scheme |
| | 2 | MMR | 2 years and older | Child incentive | £450 fine | Medical exemptions only | No compensation scheme |

* Item reversed for internal consistency check

*Block B. Order of DCE1B to DCE16B randomised (Internal Consistency check A anchored to end).*[2]

**DCE1B.** For each scenario, please consider the information from all the rows together as a whole. Compare the scenarios as if you were deciding between them in the real world. Then pick which scenario for a hypothetical mandatory vaccination scheme you prefer.

| Question number | Scenario | Vaccine | Child age group | Incentive | Penalty | Ability to opt out (exemptions) | Availability of compensation scheme |
|---|---|---|---|---|---|---|---|
| DCE1B* | 1 | MMR | 5 years and older | Parent incentive | £450 fine | Medical exemptions only | Compensation scheme |
| | 2 | 6-in-1 | 2 years and older | No incentive | Restrictions on childcare | Medical and religious beliefs | No compensation scheme |
| DCE2B | 1 | MMR | 5 years and older | Child incentive | Restrictions on childcare | Medical and religious beliefs | Compensation scheme |
| | 2 | 6-in-1 | 2 years and older | No incentive | Restrictions on Child Benefits | Medical exemptions only | No compensation scheme |
| DCE3B | 1 | MMR | 5 years and older | Child incentive | £450 fine | Medical exemptions only | No compensation scheme |
| | 2 | 6-in-1 | 2 years and older | Parent incentive | Restrictions on childcare | Medical and religious beliefs | Compensation scheme |
| DCE4B | 1 | 6-in-1 | 2 years and older | No incentive | Restrictions on Child Benefits | Medical and religious beliefs | No compensation scheme |
| | 2 | MMR | 5 years and older | Parent incentive | Restrictions on childcare | Medical exemptions only | Compensation scheme |
| DCE5B | 1 | MMR | 5 years and older | No incentive | £450 fine | Medical and religious beliefs | Compensation scheme |
| | 2 | 6-in-1 | 2 years and older | Child incentive | Restrictions on childcare | Medical exemptions only | No compensation scheme |
| DCE6B | 1 | MMR | 5 years and older | Parent incentive | Restrictions on Child Benefits | Medical exemptions only | No compensation scheme |
| | 2 | 6-in-1 | 2 years and older | Child incentive | £450 fine | Medical and religious beliefs | Compensation scheme |
| DCE7B | 1 | 6-in-1 | 5 years and older | Parent incentive | £450 fine | Medical and religious beliefs | No compensation scheme |
| | 2 | MMR | 2 years and older | No incentive | Restrictions on Child Benefits | Medical exemptions only | Compensation scheme |
| DCE8B | 1 | 6-in-1 | 2 years and older | No incentive | Restrictions on childcare | Medical exemptions only | Compensation scheme |
| | 2 | MMR | 5 years and older | Parent incentive | £450 fine | Medical and religious beliefs | No compensation scheme |
| DCE9B | 1 | 6-in-1 | 2 years and older | Child incentive | Restrictions on Child Benefits | Medical and religious beliefs | Compensation scheme |
| | 2 | MMR | 5 years and older | No incentive | £450 fine | Medical exemptions only | No compensation scheme |
| DCE10B | 1 | MMR | 5 years and older | No incentive | Restrictions on childcare | Medical and religious beliefs | Compensation scheme |
| | 2 | 6-in-1 | 2 years and older | Parent incentive | £450 fine | Medical exemptions only | No compensation scheme |
| DCE11B | 1 | MMR | 2 years and older | Parent incentive | Restrictions on Child Benefits | Medical exemptions only | Compensation scheme |

[2] DCE items displayed to participants as in example question above. For readability we have not included all items as shown to participants. Instead, we have included the combinations of attributes and levels shown to participants in each question. For exact wording of attributes and levels, see Table 1 of manuscript.

| | | | | | | | |
|---|---|---|---|---|---|---|---|
| | 2 | 6-in-1 | 5 years and older | No incentive | £450 fine | Medical and religious beliefs | No compensation scheme |
| DCE12B | 1 | MMR | 2 years and older | Parent incentive | £450 fine | Medical and religious beliefs | Compensation scheme |
| | 2 | 6-in-1 | 5 years and older | No incentive | Restrictions on childcare | Medical exemptions only | No compensation scheme |
| DCE13B | 1 | MMR | 5 years and older | Child incentive | Restrictions on Child Benefits | Medical and religious beliefs | No compensation scheme |
| | 2 | 6-in-1 | 2 years and older | Parent incentive | £450 fine | Medical exemptions only | Compensation scheme |
| DCE14B | 1 | 6-in-1 | 2 years and older | Child incentive | Restrictions on childcare | Medical exemptions only | No compensation scheme |
| | 2 | MMR | 5 years and older | No incentive | Restrictions on Child Benefits | Medical and religious beliefs | Compensation scheme |
| DCE15B | 1 | MMR | 5 years and older | No incentive | Restrictions on childcare | Medical exemptions only | No compensation scheme |
| | 2 | 6-in-1 | 2 years and older | Child incentive | Restrictions on Child Benefits | Medical and religious beliefs | Compensation scheme |
| DCE16B | 1 | 6-in-1 | 5 years and older | Parent incentive | Restrictions on childcare | Medical exemptions only | No compensation scheme |
| | 2 | MMR | 2 years and older | Child incentive | £450 fine | Medical and religious beliefs | Compensation scheme |
| Internal consistency check B | 1 | 6-in-1 | 2 years and older | No incentive | Restrictions on childcare | Medical and religious beliefs | No compensation scheme |
| | 2 | MMR | 5 years and older | Parent incentive | £450 fine | Medical exemptions only | Compensation scheme |

* Item reversed for internal consistency check

*ASK ALL*

**Q11.** To what extent, if at all, do you agree or disagree with the following statements:

SCALE:

- Strongly disagree (0) to Strongly agree (10) *[single code]*

STATEMENTS (randomise order):

- If I did not vaccinate [CHILD], [she/he] would be likely to catch the illnesses the vaccines aim to prevent
- If I did not vaccinate [CHILD], [she/he] could get severely ill from the illnesses the vaccines aim to prevent
- Child vaccinations are an effective way of preventing children from catching vaccine-preventable illnesses
- Child vaccinations are safe
- Child vaccinations cause severe side effects

**Q12.** To what extent, if at all, do you agree or disagree with the following statements:

SCALE:

- Strongly disagree (0) to Strongly agree (10). *[single code]*

STATEMENTS (randomise order):

- I approve of mandatory vaccination
- I don't like child vaccinations in general
- Vaccination campaigns are just about making money for the manufacturers
- Natural exposure to viruses and germs gives children the safest protection
- If some children do not receive vaccinations, this may cause other children to be ill with the disease
- One of my children has had a severe side effect from a routine vaccination

**Q13.** To the best of your knowledge, is [CHILD] up to date with [her/his] vaccines? *[single code]*

- [CHILD] is fully vaccinated – [she/he] has received all recommended vaccines for [her/his] age
- [CHILD] is partially vaccinated – [she/he] has received some of the recommended vaccines for [her/his] age, but not all
- [CHILD] is not vaccinated – [she/he] has not received any recommended vaccines

Finally, some questions about you and your child/children.

**Q14.** Have you ever been diagnosed by a medical doctor as having any long-lasting illness, disability or infirmity? Please select all that apply. *[multi-code]*

**You**

- Breathing complaint (e.g. asthma, bronchitis, pulmonary disease, emphysema)
- Cancer
- Diabetes
- Heart disease (e.g. heart failure, high blood pressure)
- Kidney disease (e.g. renal failure, kidney transplant)
- Liver disease (e.g. hepatitis, cirrhosis)
- Mental health (i.e. depression, anxiety, stress)
- Neurological condition (i.e. caused by disease or damage to the brain, spinal cord or other parts of the nervous system)
- Stroke (or transient ischaemic attack; TIA)
- Substance misuse (i.e. alcohol, drugs)
- Other (Please specify)
- No *[single code]*
- Don't know *[single code]*
- Prefer not to say *[single code]*

**Q15.** Has [CHILD] ever been diagnosed by a medical doctor as having any long-lasting illness, disability or infirmity? Please select all that apply. *[multi-code]*

**[CHILD]**

- Breathing complaint (e.g. asthma, bronchitis, pulmonary disease, emphysema)
- Cancer
- Diabetes
- Heart disease (e.g. heart failure, high blood pressure)
- Kidney disease (e.g. renal failure, kidney transplant)
- Liver disease (e.g. hepatitis, cirrhosis)
- Mental health (i.e. depression, anxiety, stress)
- Neurological condition (i.e. caused by disease or damage to the brain, spinal cord or other parts of the nervous system)
- Stroke (or transient ischaemic attack; TIA)
- Other (Please specify)
- No *[single code]*
- Don't know *[single code]*
- Prefer not to say *[single code]*

**Q16.** What is the highest level of educational qualification you have achieved? *[single code]*

- No qualifications
- Other qualifications (Such as NVQ level 1)
- GCSE and below (Such as O level or an RSA Diploma)
- A level or equivalent (Such as Scottish Highers or NVQ level 3)
- Higher education (Such as a HND or a NVQ level 4)
- Bachelor's Degree or equivalent (Such as a NVQ level 5)
- Master's
- PhD/Doctor
- Prefer not to say

**Q17.** What is your employment status? *[single code]*

- Full time paid job (30 hours per week or more)
- Usually working full time (30 hours per week or more), but currently furloughed
- Part time paid job (8-29 hours per week)
- Usually working part time (8-29 hours per week), but currently furloughed
- Stay-at-home parent / Looking after home / Homemaker
- Student / On a government training programme (Nation Traineeship/Modern Apprenticeship)
- Retired
- Unemployed
- Other
- Prefer not to say

**Q18.** In which of the following categories would you place your total household income from all sources before tax and any other deductions? *[single code]*

- Under £10,000
- £10,000–£19,999
- £20,000–£29,999
- £30,000–£39,999
- £40,000–£49,999
- £50,000–£74,999
- £75,000 or over
- Don't know
- Prefer not to say

**Q19.** What is your marital status? *[single code]*

- Single, never married
- Married or in a civil partnership
- Separated
- Divorced
- Widowed
- Partnered/in a relationship
- Prefer not to say

### Supplementary materials 3. Parental preferences for a mandatory vaccine scheme, sub-group analyses

Table 1 – parental education
Table 2 – total household income
Table 3 – number of children
Table 4 – ethnicity
Table 5 – region
Table 6 – partnership status
Table 7 – parent age
Table 8 – Index of Multiple Deprivation (IMD)
Table 9 – vaccine sentiment

**Table 1. Percentage of participants who have a positive preference for a mandatory vaccine scheme attribute, by parental education.**

Table 1 (A). Participants who did not attend university (n=553).

| Attribute | Level | Mean (standard error) | % who prefer this level (95% CI) | *p*-level | Relative importance |
|---|---|---|---|---|---|
| Vaccine | MMR | | Reference | | 12.9% |
| | 6-in-1 | 0.16 (0.04) | 59.5 (54.8 to 64.2) | <0.001 | |
| Child age group | 2 years and older | | Reference | | 9.2% |
| | 5 years and older | -0.11 (0.04) | 43.8 (39.3 to 48.3) | 0.007 | |
| Incentive | No incentive | | Reference | | 28.5% |
| | Incentive for child (£130 voucher) | 0.30 (0.04) | 77.5 (71.0 to 83.0) | <0.001 | |
| | Incentive for parent (£130 cash pay-out) | 0.34 (0.05) | 70.0 (65.0 to 74.7) | <0.001 | |
| Penalty | £450 fine | | Reference | | 31.3% |
| | Child not able to attend school or childcare / day care until they are vaccinated | 0.38 (0.05) | 71.9 (67.0 to 76.3) | <0.001 | |
| | Parent not able to claim Child Benefits for child until they are vaccinated | 0.26 (0.05) | 65.5 (60.1 to 70.6) | <0.001 | |
| Ability to opt out | Medical exemption | | Reference | | 0.9% |
| | Medical and religious belief exemption | 0.01 (0.03) | 51.8 (42.2 to 61.2) | 0.72 | |
| Compensation scheme | Not offered | | Reference | | 17.2% |
| | Offered | 0.21 (0.04) | 65.8 (60.7 to 70.5) | <0.001 | |

Log likelihood = -5724.95, Likelihood Ratio Test=5256.51, Akaike's Information Criterion=11483.9

Table 1 (B). Participants educated to degree level or higher (n=442).

| Attribute | Level | Mean (standard error) | % who prefer this level (95% CI) | *p*-level | Relative importance |
|---|---|---|---|---|---|
| Vaccine | MMR | | Reference | | 7.1% |
| | 6-in-1 | 0.10 (0.05) | 56.5 (50.9 to 62.0) | 0.02 | |
| Child age group | 2 years and older | | Reference | | 12.0% |
| | 5 years and older | -0.19 (0.05) | 41.4 (36.8 to 46.0) | <0.001 | |
| Incentive | No incentive | | Reference | | 31.4% |
| | Incentive for child (£130 voucher) | 0.37 (0.05) | 87.8 (81.2 to 92.6) | <0.001 | |
| | Incentive for parent (£130 cash pay-out) | 0.45 (0.05) | 82.7 (77.0 to 87.4) | <0.001 | |
| Penalty | £450 fine | | Reference | | 22.9% |
| | Child not able to attend school or childcare / day care until they are vaccinated | 0.27 (0.05) | 66.6 (60.6 to 72.2) | <0.001 | |
| | Parent not able to claim Child Benefits for child until they are vaccinated | 0.33 (0.05) | 70.3 (64.2 to 75.9) | <0.001 | |
| Ability to opt out | Medical exemption | | Reference | | 8.6% |
| | Medical and religious belief exemption | -0.14 (0.04) | 41.1 (35.8 to 46.5) | 0.001 | |
| Compensation scheme | Not offered | | Reference | | 18.0% |
| | Offered | 0.27 (0.04) | 68.4 (63.0 to 73.3) | <0.001 | |

Log likelihood = -4493, Likelihood Ratio Test=539.73, Akaike's Information Criterion=9020.06

**Table 2. Percentage of participants who have a positive preference for the mandatory vaccine scheme attribute, by total household income**

Table 2 (A). Participants with a lower household income (total annual income of up to £29,999; n=352).

| Attribute | Level | Mean (standard error) | % who prefer this level (95% CI) | *p*-level | Relative importance |
|---|---|---|---|---|---|
| Vaccine | MMR | | Reference | | 10.0% |
| | 6-in-1 | 0.10 (0.04) | 59.4 (51.7 to 66.8) | 0.02 | |
| Child age group | 2 years and older | | Reference | | 9.2% |
| | 5 years and older | -0.09 (0.05) | 44.9 (39.4 to 50.4) | 0.07 | |
| Incentive | No incentive | | Reference | | 38.0% |
| | Incentive for child (£130 voucher) | 0.31 (0.05) | 91.6 (83.6 to 96.2) | <0.001 | |
| | Incentive for parent (£130 cash pay-out) | 0.37 (0.05) | 77.0 (70.3 to 82.8) | <0.001 | |
| Penalty | £450 fine | | Reference | | 28.0% |
| | Child not able to attend school or childcare / day care until they are vaccinated | 0.27 (0.05) | 75.0 (67.0 to 81.8) | <0.001 | |
| | Parent not able to claim Child Benefits for child until they are vaccinated | 0.15 (0.05) | 64.1 (54.8 to 72.7) | 0.003 | |
| Ability to opt out | Medical exemption | | Reference | | 0.4% |
| | Medical and religious belief exemption | 0.00 (0.04) | 50.5 (41.6 to 59.3) | 0.92 | |
| Compensation scheme | Not offered | | Reference | | 14.5% |
| | Offered | 0.14 (0.04) | 63.4 (56.2 to 70.1) | <0.001 | |

Log likelihood =-3703.99, Likelihood Ratio Test=227.33, Akaike's Information Criterion=7566.58

Table 2 (B). Participants with a higher household income (total annual income of £30,000 or over per year, n=618).

| Attribute | Level | Mean (standard error) | % who prefer this level (95% CI) | *p*-level | Relative importance |
|---|---|---|---|---|---|
| Vaccine | MMR | | Reference | | 10.8% |
| | 6-in-1 | 0.15 (0.04) | 57.7 (53.4 to 61.9) | <0.001 | |
| Child age group | 2 years and older | | Reference | | 11.8% |
| | 5 years and older | -0.15 (0.04) | 43.0 (39.0 to 47.1) | 0.001 | |
| Incentive | No incentive | | Reference | | 27.4% |
| | Incentive for child (£130 voucher) | 0.33 (0.04) | 80.4 (74.4 to 85.5) | <0.001 | |
| | Incentive for parent (£130 cash pay-out) | 0.39 (0.05) | 73.3 (68.5 to 77.8) | <0.001 | |
| Penalty | £450 fine | | Reference | | 25.2% |
| | Parent not able to claim Child Benefits for child until they are vaccinated | 0.33 (0.05) | 66.9 (62.3 to 71.3) | <0.001 | |
| | Child not able to attend school or childcare / day care until they are vaccinated | 0.37 (0.05) | 68.2 (63.6 to 72.5) | <0.001 | |
| Ability to opt out | Medical exemption | | Reference | | 5.6% |
| | Medical and religious belief exemption | -0.08 (0.04) | 43.7 (38.7 to 48.9) | 0.02 | |
| Compensation scheme | Not offered | | Reference | | 19.2% |
| | Offered | 0.29 (0.04) | 68.1 (63.8 to 72.1) | <0.001 | |

Log likelihood =-6262.90, Likelihood Ratio Test=820.34, Akaike's Information Criterion=12559.8

**Table 3. Percentage of participants who have a positive preference for the mandatory vaccine scheme attribute, by number of children**

Table 3 (A). Participants with one child (n=365).

| Attribute | Level | Mean (standard error) | % who prefer this level (95% CI) | *p*-level | Relative importance |
|---|---|---|---|---|---|
| Vaccine | MMR | | Reference | | 10.0% |
| | 6-in-1 | 0.12 (0.05) | 57.5 (51.5 to 63.3) | 0.01 | |
| Child age group | 2 years and older | | Reference | | 12.5% |
| | 5 years and older | -0.16 (0.06) | 42.6 (37.5 to 47.9) | 0.006 | |
| Incentive | No incentive | | Reference | | 29.2% |
| | Incentive for child (£130 voucher) | 0.25 (0.05) | 84.0 (73.4 to 91.3) | <0.001 | |
| | Incentive for parent (£130 cash pay-out) | 0.36 (0.05) | 79.6 (72.3 to 85.5) | <0.001 | |
| Penalty | £450 fine | | Reference | | 22.6% |
| | Child not able to attend school or childcare / day care until they are vaccinated | 0.23 (0.06) | 64.5 (57.9 to 70.8) | <0.001 | |
| | Parent not able to claim Child Benefits for child until they are vaccinated | 0.28 (0.06) | 67.7 (60.8 to 73.9) | <0.001 | |
| Ability to opt out | Medical exemption | | Reference | | 6.4% |
| | Medical and religious belief exemption | -0.08 (0.04) | 39.8 (30.7 to 49.4) | 0.04 | |
| Compensation scheme | Not offered | | Reference | | 19.3% |
| | Offered | 0.24 (0.05) | 66.1 (60.3 to 71.6) | <0.001 | |

Log likelihood =3742.70, Likelihood Ratio Test=425.85, Akaike's Information Criterion=7519.40

Table 3 (B). Participants with two children (n=391).

| Attribute | Level | Mean (standard error) | % who prefer this level (95% CI) | *p*-level | Relative importance |
|---|---|---|---|---|---|
| Vaccine | MMR | | Reference | | 10.9% |
| | 6-in-1 | 0.15 (0.05) | 59.5 (53.5 to 65.3) | 0.002 | |
| Child age group | 2 years and older | | Reference | | 13.5% |
| | 5 years and older | -0.19 (0.05) | 40.7 (35.6 to 45.9) | <0.001 | |
| Incentive | No incentive | | Reference | | 26.7% |
| | Incentive for child (£130 voucher) | 0.31 (0.05) | 84.2 (75.7 to 90.4) | <0.001 | |
| | Incentive for parent (£130 cash pay-out) | 0.37 (0.06) | 72.1 (65.9 to 77.7) | <0.001 | |
| Penalty | £450 fine | | Reference | | 29.5% |
| | Parent not able to claim Child Benefits for child until they are vaccinated | 0.41 (0.06) | 71.9 (66.2 to 77.0) | <0.001 | |
| | Child not able to attend school or childcare / day care until they are vaccinated | 0.35 (0.06) | 69.0 (62.9 to 74.6) | <0.001 | |
| Ability to opt out | Medical exemption | | Reference | | 4.2% |
| | Medical and religious belief exemption | -0.06 (0.04) | 45.7 (39.5 to 52.1) | 0.19 | |
| Compensation scheme | Not offered | | Reference | | 15.2% |
| | Offered | 0.21 (0.05) | 63.8 (58.1 to 69.2) | <0.001 | |

Log likelihood =3988.31, Likelihood Ratio Test=450.56, Akaike's Information Criterion=8010.63

Table 3 (C). Participants with three or more children (n=245).

| Attribute | Level | Mean (standard error) | % who prefer this level (95% CI) | *p*-level | Relative importance |
|---|---|---|---|---|---|
| Vaccine | MMR | | Reference | | 11.8% |
| | 6-in-1 | 0.13 (0.06) | 59.1 (51.5 to 66.4) | 0.02 | |
| Child age group | 2 years and older | | Reference | | 1.6% |
| | 5 years and older | -0.02 (0.06) | 48.8 (41.5 to 56.1) | 0.74 | |
| Incentive | No incentive | | Reference | | 39.1% |
| | Incentive for child (£130 voucher) | 0.44 (0.06) | 87.6 (80.1 to 92.9) | <0.001 | |
| | Incentive for parent (£130 cash pay-out) | 0.44 (0.07) | 76.4 (69.1 to 82.6) | <0.001 | |
| Penalty | £450 fine | | Reference | | 28.4% |
| | Child not able to attend school or childcare / day care until they are vaccinated | 0.32 (0.06) | 72.3 (64.1 to 79.5) | <0.001 | |
| | Parent not able to claim Child Benefits for child until they are vaccinated | 0.22 (0.07) | 66.2 (56.9 to 74.6) | 0.001 | |
| Ability to opt out | Medical exemption | | Reference | | 0.6% |
| | Medical and religious belief exemption | 0.01 (0.05) | 50.7 (41.2 to 60.1) | 0.89 | |
| Compensation scheme | Not offered | | Reference | | 18.5% |
| | Offered | 0.21 (0.04) | 74.8 (64.9 to 82.9) | <0.001 | |

Log likelihood =2563.51, Likelihood Ratio Test=161.44, Akaike's Information Criterion=5161.02

**Table 4. Percentage of participants who have a positive preference for the mandatory vaccine scheme attribute, by ethnicity**

Table 4 (A). Participants of white ethnicity (n=839).

| Attribute | Level | Mean (standard error) | % who prefer this level (95% CI) | *p*-level | Relative importance |
|---|---|---|---|---|---|
| Vaccine | MMR | | Reference | | 13.6% |
| | 6-in-1 | 0.18 (0.03) | 60.8 (56.8 to 64.7) | <0.001 | |
| Child age group | 2 years and older | | Reference | | 11.3% |
| | 5 years and older | -0.15 (0.04) | 42.7 (39.2 to 46.2) | <0.001 | |
| Incentive | No incentive | | Reference | | 29.9% |
| | Incentive for child (£130 voucher) | 0.33 (0.03) | 82.9 (77.8 to 87.2) | <0.001 | |
| | Incentive for parent (£130 cash pay-out) | 0.39 (0.04) | 73.9 (69.8 to 77.7) | <0.001 | |
| Penalty | £450 fine | | Reference | | 24.8% |
| | Child not able to attend school or childcare / day care until they are vaccinated | 0.32 (0.04) | 68.7 (64.6 to 72.6) | <0.001 | |
| | Parent not able to claim Child Benefits for child until they are vaccinated | 0.28 (0.04) | 66.5 (62.2 to 70.6) | <0.001 | |
| Ability to opt out | Medical exemption | | Reference | | 3.0% |
| | Medical and religious belief exemption | -0.04 (0.03) | 46.7 (42.0 to 51.4) | 0.17 | |
| Compensation scheme | Not offered | | Reference | | 17.3% |
| | Offered | 0.22 (0.03) | 65.9 (61.9 to 69.7) | <0.001 | |

Log likelihood =-815.67, Likelihood Ratio Test=911.29, Akaike's Information Criterion=17265.33

Table 4 (B). Participants from black and minoritised ethnic groups (n=151).

| Attribute | Level | Mean (standard error) | % who prefer this level (95% CI) | *p*-level | Relative importance |
|---|---|---|---|---|---|
| Vaccine | MMR | | Reference | | 1.8% |
| | 6-in-1 | -0.02 (0.07) | 48.4 (38.3 to 58.5) | 0.75 | |
| Child age group | 2 years and older | | Reference | | 5.7% |
| | 5 years and older | -0.07 (0.07) | 45.7 (36.9 to 54.8) | 0.36 | |
| Incentive | No incentive | | Reference | | 29.7% |
| | Incentive for child (£130 voucher) | 0.26 (0.07) | 89.0 (72.5 to 96.8) | <0.001 | |
| | Incentive for parent (£130 cash pay-out) | 0.34 (0.08) | 80.5 (68.6 to 89.2) | <0.001 | |
| Penalty | £450 fine | | Reference | | 28.8% |
| | Child not able to attend school or childcare / day care until they are vaccinated | 0.33 (0.07) | 81.1 (69.2 to 89.6) | <0.001 | |
| | Parent not able to claim Child Benefits for child until they are vaccinated | 0.28 (0.08) | 74.9 (62.0 to 85.0) | <0.001 | |
| Ability to opt out | Medical exemption | | Reference | | 11.1% |
| | Medical and religious belief exemption | -0.13 (0.05) | 27.6 (13.7 to 46.1) | 0.02 | |
| Compensation scheme | Not offered | | Reference | | 22.9% |
| | Offered | 0.27 (0.06) | 73.2 (63.4 to 81.5) | <0.001 | |

Log likelihood =-1581.39, Likelihood Ratio Test=94.15, Akaike's Information Criterion=3196.78

**Table 5. Percentage of participants who have a positive preference for the mandatory vaccine scheme attribute, by region**

Table 5 (A). Participants living in London (n=183).

| Attribute | Level | Mean (standard error) | % who prefer this level (95% CI) | p-level | Relative importance |
|---|---|---|---|---|---|
| Vaccine | MMR | | Reference | | 13.1% |
| | 6-in-1 | 0.13 (0.07) | 57.4 (49.5 to 65.1) | 0.07 | |
| Child age group | 2 years and older | | Reference | | 2.0% |
| | 5 years and older | -0.02 (0.07) | 48.9 (40.9 to 56.8) | 0.78 | |
| Incentive | No incentive | | Reference | | 26.2% |
| | Incentive for child (£130 voucher) | 0.25 (0.06) | 100.0 (99.7 to 100.0)& | <0.001 | |
| | Incentive for parent (£130 cash pay-out) | 0.25 (0.07) | 72.2 (60.4 to 82.0) | <0.001 | |
| Penalty | £450 fine | | Reference | | 30.5% |
| | Child not able to attend school or childcare / day care until they are vaccinated | 0.20 (0.07) | 66.7 (55.6 to 76.5) | 0.004 | |
| | Parent not able to claim Child Benefits for child until they are vaccinated | 0.30 (0.07) | 71.8 (61.5 to 80.5) | <0.001 | |
| Ability to opt out | Medical exemption | | Reference | | 12.1% |
| | Medical and religious belief exemption | -0.12 (0.05) | 36.6 (25.7 to 48.7) | 0.03 | |
| Compensation scheme | Not offered | | Reference | | 16.2% |
| | Offered | 0.16 (0.05) | 70.0 (57.7 to 80.4) | 0.002 | |

Log likelihood =1922.37, Likelihood Ratio Test=150.99, Akaike's Information Criterion=3878.75.

[&]Model parameter unreliable due to a small sample size

Table 5 (B). Participants living in the South (South East, South West; n=224).

| Attribute | Level | Mean (standard error) | % who prefer this level (95% CI) | p-level | Relative importance |
|---|---|---|---|---|---|
| Vaccine | MMR | | Reference | | 13.4% |
| | 6-in-1 | 0.21 (0.07) | 62.1 (54.5 to 69.2) | 0.002 | |
| Child age group | 2 years and older | | Reference | | 13.5% |
| | 5 years and older | -0.21 (0.08) | 40.6 (34.3 to 47.1) | 0.005 | |
| Incentive | No incentive | | Reference | | 21.2% |
| | Incentive for child (£130 voucher) | 0.32 (0.07) | 76.7 (66.4 to 84.9) | <0.001 | |
| | Incentive for parent (£130 cash pay-out) | 0.34 (0.07) | 74.5 (65.1 to 82.4) | <0.001 | |
| Penalty | £450 fine | | Reference | | 17.2% |
| | Child not able to attend school or childcare / day care until they are vaccinated | 0.25 (0.07) | 66.5 (57.5 to 74.6) | <0.001 | |
| | Parent not able to claim Child Benefits for child until they are vaccinated | 0.27 (0.08) | 65.5 (56.9 to 73.3) | 0.001 | |
| Ability to opt out | Medical exemption | | Reference | | 13.4% |
| | Medical and religious belief exemption | -0.21 (0.07) | 38.0 (31.1 to 45.2) | 0.001 | |
| Compensation scheme | Not offered | | Reference | | 21.3% |
| | Offered | 0.34 (0.06) | 71.1 (64.0 to 77.5) | <0.001 | |

Log likelihood =2550.31, Likelihood Ratio Test=308.85, Akaike's Information Criterion=4534.62

Table 5 (C). Participants living in the midlands (East Midlands, West Midlands, East of England; n=305).

| Attribute | Level | Mean (standard error) | % who prefer this level (95% CI) | *p*-level | Relative importance |
|---|---|---|---|---|---|
| Vaccine | MMR | | Reference | | 6.0% |
| | 6-in-1 | 0.07 (0.05) | 55.0 (47.5 to 62.3) | 0.19 | |
| Child age group | 2 years and older | | Reference | | 9.6% |
| | 5 years and older | -0.10 (0.06) | 44.9 (39.2 to 50.7) | 0.08 | |
| Incentive | No incentive | | Reference | | 32.6% |
| | Incentive for child (£130 voucher) | 0.29 (0.06) | 74.0 (65.5 to 81.3) | <0.001 | |
| | Incentive for parent (£130 cash pay-out) | 0.36 (0.06) | 71.0 (64.1 to 77.2) | <0.001 | |
| Penalty | £450 fine | | Reference | | 29.1% |
| | Child not able to attend school or childcare / day care until they are vaccinated | 0.32 (0.06) | 68.4 (61.6 to 74.7) | <0.001 | |
| | Parent not able to claim Child Benefits for child until they are vaccinated | 0.21 (0.06) | 66.4 (57.5 to 74.3) | <0.001 | |
| Ability to opt out | Medical exemption | | Reference | | 0.7% |
| | Medical and religious belief exemption | -0.01 (0.04) | 49.2 (40.6 to 57.9) | 0.86 | |
| Compensation scheme | Not offered | | Reference | | 21.9% |
| | Offered | 0.24 (0.05) | 65.9 (59.5 to 71.8) | <0.001 | |

Log likelihood =-3155.45, Likelihood Ratio Test=312.31, Akaike's Information Criterion=6344.90

Table 5 (D). Participants living in the North (North East, North West, Yorkshire and the Humber; n=289).

| Attribute | Level | Mean (standard error) | % who prefer this level (95% CI) | *p*-level | Relative importance |
|---|---|---|---|---|---|
| Vaccine | MMR | | Reference | | 10.0% |
| | 6-in-1 | 0.18 (0.06) | 60.8 (54.1 to 67.2) | 0.002 | |
| Child age group | 2 years and older | | Reference | | 11.5% |
| | 5 years and older | -0.19 (0.06) | 39.5 (33.4 to 45.9) | 0.001 | |
| Incentive | No incentive | | Reference | | 34.3% |
| | Incentive for child (£130 voucher) | 0.42 (0.06) | 86.4 (78.9 to 91.8) | <0.001 | |
| | Incentive for parent (£130 cash pay-out) | 0.56 (0.07) | 80.1 (74.0 to 82.5) | <0.001 | |
| Penalty | £450 fine | | Reference | | 26.4% |
| | Child not able to attend school or childcare / day care until they are vaccinated | 0.43 (0.07) | 72.8 (66.4 to 78.5) | <0.001 | |
| | Parent not able to claim Child Benefits for child until they are vaccinated | 0.39 (0.07) | 68.3 (61.8 to 74.2) | 0.001 | |
| Ability to opt out | Medical exemption | | Reference | | 4.6% |
| | Medical and religious belief exemption | 0.08 (0.04) | 60.6 (48.6 to 71.7) | 0.001 | |
| Compensation scheme | Not offered | | Reference | | 12.3% |
| | Offered | 0.20 (0.05) | 63.2 (56.6 to 69.5) | <0.001 | |

Log likelihood =-2933.18, Likelihood Ratio Test=303.57, Akaike's Information Criterion=5900.36

**Table 6. Percentage of participants who have a positive preference for the mandatory vaccine scheme attribute, by partnership status**

Table 6 (A). Participants who were not partnered (n=169).

| Attribute | Level | Mean (standard error) | % who prefer this level (95% CI) | *p*-level | Relative importance |
|---|---|---|---|---|---|
| Vaccine | MMR | | Reference | | 12.5% |
| | 6-in-1 | 0.15 (0.06) | 62.0 (51.9 to 71.4) | 0.02 | |
| Child age group | 2 years and older | | Reference | | 0.8% |
| | 5 years and older | -0.01 (0.08) | 49.5 (41.9 to 57.2) | 0.90 | |
| Incentive | No incentive | | Reference | | 36.2% |
| | Incentive for child (£130 voucher) | 0.31 (0.07) | 93.5 (81.3 to 98.4) | <0.001 | |
| | Incentive for parent (£130 cash pay-out) | 0.42 (0.07) | 87.7 (78.3 to 93.9) | <0.001 | |
| Penalty | £450 fine | | Reference | | 27.1% |
| | Child not able to attend school or childcare / day care until they are vaccinated | 0.32 (0.07) | 76.6 (65.8 to 85.2) | <0.001 | |
| | Parent not able to claim Child Benefits for child until they are vaccinated | 0.28 (0.07) | 77.1 (64.1 to 86.9) | <0.001 | |
| Ability to opt out | Medical exemption | | Reference | | 10.9% |
| | Medical and religious belief exemption | 0.13 (0.05) | 69.1 (53.9 to 81.5) | 0.01 | |
| Compensation scheme | Not offered | | Reference | | 12.4% |
| | Offered | 0.14 (0.06) | 65.7 (54.1 to 76.0) | 0.008 | |

Log likelihood =-1758.19, Likelihood Ratio Test=139.60, Akaike's Information Criterion=3550.37

Table 6 (B). Partnered participants (n=830).

| Attribute | Level | Mean (standard error) | % who prefer this level (95% CI) | *p*-level | Relative importance |
|---|---|---|---|---|---|
| Vaccine | MMR | | Reference | | 10.2% |
| | 6-in-1 | 0.14 (0.03) | 58.0 (54.2 to 61.8) | <0.001 | |
| Child age group | 2 years and older | | Reference | | 12.4% |
| | 5 years and older | -0.17 (0.04) | 41.4 (37.9 to 45.0) | <0.001 | |
| Incentive | No incentive | | Reference | | 28.4% |
| | Incentive for child (£130 voucher) | 0.34 (0.03) | 81.1 (76.0 to 85.4) | <0.001 | |
| | Incentive for parent (£130 cash pay-out) | 0.38 (0.04) | 73.0 (68.8 to 76.8) | <0.001 | |
| Penalty | £450 fine | | Reference | | 23.5% |
| | Child not able to attend school or childcare / day care until they are vaccinated | 0.32 (0.04) | 67.8 (63.7 to 71.7) | <0.001 | |
| | Parent not able to claim Child Benefits for child until they are vaccinated | 0.29 (0.04) | 66.2 (62.0 to 70.3) | <0.001 | |
| Ability to opt out | Medical exemption | | Reference | | 6.9% |
| | Medical and religious belief exemption | -0.09 (0.03) | 42.3 (37.7 to 47.0) | 0.001 | |
| Compensation scheme | Not offered | | Reference | | 18.6% |
| | Offered | 0.25 (0.03) | 66.5 (62.7 to 70.1) | <0.001 | |

Log likelihood =-8510.08, Likelihood Ratio Test=910.31, Akaike's Information Criterion=17054.16

**Table 7. Percentage of participants who have a positive preference for the mandatory vaccine scheme attribute, by parent age**

Table 7 (A). Participants aged under 30 years (n=280).

| Attribute | Level | Mean (standard error) | % who prefer this level (95% CI) | *p*-level | Relative importance |
|---|---|---|---|---|---|
| Vaccine | MMR | | Reference | | 9.8% |
| | 6-in-1 | 0.08 (0.04) | 58.7 (49.4 to 67.6) | 0.07 | |
| Child age group | 2 years and older | | Reference | | 10.4% |
| | 5 years and older | -0.08 (0.05) | 44.1 (37.5 to 50.9) | 0.09 | |
| Incentive | No incentive | | Reference | | 33.2% |
| | Incentive for child (£130 voucher) | 0.21 (0.05) | 89.8 (76.0 to 96.7) | <0.001 | |
| | Incentive for parent (£130 cash pay-out) | 0.27 (0.06) | 73.3 (64.4 to 80.9) | <0.001 | |
| Penalty | £450 fine | | Reference | | 19.1% |
| | Child not able to attend school or childcare / day care until they are vaccinated | 0.15 (0.05) | 64.3 (54.7 to 73.0) | 0.004 | |
| | Parent not able to claim Child Benefits for child until they are vaccinated | 0.08 (0.05) | 57.9 (47.0 to 68.2) | 0.16 | |
| Ability to opt out | Medical exemption | | Reference | | 5.7% |
| | Medical and religious belief exemption | 0.05 (0.04) | 57.5 (44.8 to 69.5) | 0.25 | |
| Compensation scheme | Not offered | | Reference | | 21.8% |
| | Offered | 0.18 (0.04) | 66.3 (58.7 to 73.3) | <0.001 | |

Log likelihood =-2998.17, Likelihood Ratio Test=121.49, Akaike's Information Criterion=6030.27

Table 7 (B). Participants aged 30 years or over (n=721).

| Attribute | Level | Mean (standard error) | % who prefer this level (95% CI) | *p*-level | Relative importance |
|---|---|---|---|---|---|
| Vaccine | MMR | | Reference | | 11.2% |
| | 6-in-1 | 0.17 (0.04) | 58.8 (54.8 to 62.8) | <0.001 | |
| Child age group | 2 years and older | | Reference | | 9.8% |
| | 5 years and older | -0.15 (0.04) | 43.3 (39.6 to 47.0) | <0.001 | |
| Incentive | No incentive | | Reference | | 28.5% |
| | Incentive for child (£130 voucher) | 0.39 (0.04) | 82.9 (77.9 to 87.1) | <0.001 | |
| | Incentive for parent (£130 cash pay-out) | 0.43 (0.04) | 74.7 (70.4 to 78.6) | <0.001 | |
| Penalty | £450 fine | | Reference | | 26.7% |
| | Child not able to attend school or childcare / day care until they are vaccinated | 0.40 (0.04) | 71.0 (66.8 to 74.9) | <0.001 | |
| | Parent not able to claim Child Benefits for child until they are vaccinated | 0.38 (0.04) | 70.3 (65.9 to 74.3) | <0.001 | |
| Ability to opt out | Medical exemption | | Reference | | 6.3% |
| | Medical and religious belief exemption | -0.09 (0.03) | 42.5 (37.6 to 47.4) | 0.003 | |
| Compensation scheme | Not offered | | Reference | | 17.5% |
| | Offered | 0.26 (0.03) | 67.0 (62.9 to 70.9) | <0.001 | |

Log likelihood =-7278.73, Likelihood Ratio Test=949.15, Akaike's Information Criterion=14591.13

**Table 8. Percentage of participants who have a positive preference for the mandatory vaccine scheme attribute, by Index of Multiple Deprivation**

Table 8 (A). Participants who live in the most deprived areas (deciles 1 to 3; n=383).

| Attribute | Level | Mean (standard error) | % who prefer this level (95% CI) | *p*-level | Relative importance |
|---|---|---|---|---|---|
| Vaccine | MMR | | Reference | | 14.8% |
| | 6-in-1 | 0.15 (0.04) | 63.3 (56.3 to 69.9) | <0.001 | |
| Child age group | 2 years and older | | Reference | | 7.9% |
| | 5 years and older | -0.08 (0.05) | 45.4 (40.0 to 50.8) | 0.10 | |
| Incentive | No incentive | | Reference | | 33.0% |
| | Incentive for child (£130 voucher) | 0.27 (0.04) | 81.6 (72.9 to 88.4) | <0.001 | |
| | Incentive for parent (£130 cash pay-out) | 0.33 (0.05) | 76.6 (69.6 to 82.5) | <0.001 | |
| Penalty | £450 fine | | Reference | | 24.5% |
| | Child not able to attend school or childcare / day care until they are vaccinated | 0.24 (0.05) | 67.6 (60.9 to 73.8) | <0.001 | |
| | Parent not able to claim Child Benefits for child until they are vaccinated | 0.17 (0.05) | 61.5 (54.6 to 68.0) | 0.001 | |
| Ability to opt out | Medical exemption | | Reference | | 2.7% |
| | Medical and religious belief exemption | 0.03 (0.03) | 53.9 (43.8 to 63.8) | 0.45 | |
| Compensation scheme | Not offered | | Reference | | 17.1% |
| | Offered | 0.17 (0.04) | 70.7 (62.8 to 77.8) | <0.001 | |

Log likelihood =-4040.36, Likelihood Ratio Test=244.19, Akaike's Information Criterion=8586.44

Table 8 (B). Participants who live in neither the least nor the most deprived areas (deciles 4 to 7; n=420).

| Attribute | Level | Mean (standard error) | % who prefer this level (95% CI) | *p*-level | Relative importance |
|---|---|---|---|---|---|
| Vaccine | MMR | | Reference | | 8.3% |
| | 6-in-1 | 0.12 (0.05) | 56.2 (51.0 to 61.3) | 0.02 | |
| Child age group | 2 years and older | | Reference | | 12.0% |
| | 5 years and older | -0.17 (0.05) | 41.0 (36.1 to 46.1) | 0.001 | |
| Incentive | No incentive | | Reference | | 32.4% |
| | Incentive for child (£130 voucher) | 0.37 (0.05) | 86.9 (80.1 to 91.9) | <0.001 | |
| | Incentive for parent (£130 cash pay-out) | 0.46 (0.06) | 76.2 (70.7 to 81.1) | <0.001 | |
| Penalty | £450 fine | | Reference | | 25.3% |
| | Child not able to attend school or childcare / day care until they are vaccinated | 0.35 (0.06) | 68.8 (63.1 to 74.0) | <0.001 | |
| | Parent not able to claim Child Benefits for child until they are vaccinated | 0.36 (0.06) | 71.6 (65.5 to 77.1) | <0.001 | |
| Ability to opt out | Medical exemption | | Reference | | 6.3% |
| | Medical and religious belief exemption | -0.09 (0.04) | 42.3 (35.7 to 49.2) | 0.03 | |
| Compensation scheme | Not offered | | Reference | | 15.7% |
| | Offered | 0.22 (0.05) | 63.6 (58.3 to 68.7) | <0.001 | |

Log likelihood =-4276.21, Likelihood Ratio Test=499.38, Akaike's Information Criterion=8586.44

Table 8 (C). Participants who live in the least deprived areas (deciles 8 to 10; n=198).

| Attribute | Level | Mean (standard error) | % who prefer this level (95% CI) | *p*-level | Relative importance |
|---|---|---|---|---|---|
| Vaccine | MMR | | Reference | | 7.0% |
| | 6-in-1 | 0.12 (0.08) | 56.4 (48.4 to 64.1) | 0.12 | |
| Child age group | 2 years and older | | Reference | | 9.8% |
| | 5 years and older | -0.17 (0.10) | 44.0 (37.4 to 50.8) | 0.08 | |
| Incentive | No incentive | | Reference | | 22.0% |
| | Incentive for child (£130 voucher) | 0.37 (0.08) | 75.5 (65.6 to 83.6) | <0.001 | |
| | Incentive for parent (£130 cash pay-out) | 0.38 (0.09) | 70.0 (61.3 to 77.7) | <0.001 | |
| Penalty | £450 fine | | Reference | | 28.0% |
| | Child not able to attend school or childcare / day care until they are vaccinated | 0.48 (0.09) | 72.7 (64.9 to 79.5) | <0.001 | |
| | Parent not able to claim Child Benefits for child until they are vaccinated | 0.41 (0.09) | 68.8 (60.7 to 76.1) | <0.001 | |
| Ability to opt out | Medical exemption | | Reference | | 9.5% |
| | Medical and religious belief exemption | -0.16 (0.07) | 40.3 (32.6 to 48.4) | 0.02 | |
| Compensation scheme | Not offered | | Reference | | 23.7% |
| | Offered | 0.41 (0.08) | 68.7 (62.1 to 74.8) | <0.001 | |

Log likelihood =-1944.61, Likelihood Ratio Test=367.88, Akaike's Information Criterion=3923.23

**Table 9. Percentage of participants who have a positive preference for the mandatory vaccine scheme attribute, by vaccine sentiment**

Table 9 (A). Participants with the least positive vaccine sentiments (n=330).

| Attribute | Level | Mean (standard error) | % who prefer this level (95% CI) | *p*-level | Relative importance |
|---|---|---|---|---|---|
| Vaccine | MMR | | Reference | | 7.6% |
| | 6-in-1 | 0.08 (0.04) | 57.0 (49.6 to 64.1) | 0.06 | |
| Child age group | 2 years and older | | Reference | | 9.3% |
| | 5 years and older | 0.10 (0.05) | 56.2 (50.2 to 62.0) | 0.04 | |
| Incentive | No incentive | | Reference | | |
| | Incentive for child (£130 voucher) | 0.20 (0.05) | 81.1 (68.0 to 90.2) | <0.001 | 24.8% |
| | Incentive for parent (£130 cash pay-out) | 0.27 (0.05) | 70.5 (62.8 to 77.4) | <0.001 | |
| Penalty | £450 fine | | Reference | | |
| | Child not able to attend school or childcare / day care until they are vaccinated | 0.16 (0.05) | 63.9 (55.3 to 71.8) | 0.002 | 28.7% |
| | Parent not able to claim Child Benefits for child until they are vaccinated | 0.31 (0.06) | 72.6 (65.1 to 79.1) | <0.001 | |
| Ability to opt out | Medical exemption | | Reference | | 8.2% |
| | Medical and religious belief exemption | 0.09 (0.04) | 60.2 (51.2 to 68.6) | 0.03 | |
| Compensation scheme | Not offered | | Reference | | 21.3% |
| | Offered | 0.23 (0.04) | 69.2 (62.6 to 75.3) | <0.001 | |

Log likelihood =-3488.09, Likelihood Ratio Test=216.70, Akaike's Information Criterion=7010.18

Table 9 (B). Participants scoring in the middle range for positive vaccine sentiments (n=337).

| Attribute | Level | Mean (standard error) | % who prefer this level (95% CI) | *p*-level | Relative importance |
|---|---|---|---|---|---|
| Vaccine | MMR | | Reference | | 4.2% |
| | 6-in-1 | 0.06 (0.05) | 54.3 (47.4 to 61.0) | 0.22 | |
| Child age group | 2 years and older | | Reference | | 15.8% |
| | 5 years and older | -0.22 (0.05) | 37.4 (32.0 to 43.2) | . <0.001 | |
| Incentive | No incentive | | Reference | | |
| | Incentive for child (£130 voucher) | 0.38 (0.05) | 89.1 (82.1 to 93.9) | <0.001 | 29.0% |
| | Incentive for parent (£130 cash pay-out) | 0.40 (0.06) | 77.6 (70.9 to 83.3) | <0.001 | |
| Penalty | £450 fine | | Reference | | |
| | Child not able to attend school or childcare / day care until they are vaccinated | 0.41 (0.06) | 75.1 (68.9 to 80.6) | <0.001 | 30.0% |
| | Parent not able to claim Child Benefits for child until they are vaccinated | 0.34 (0.06) | 73.4 (66.3 to 79.7) | <0.001 | |
| Ability to opt out | Medical exemption | | Reference | | 7.1% |
| | Medical and religious belief exemption | -0.10 (0.04) | 39.9 (31.9 to 48.3) | 0.02 | |
| Compensation scheme | Not offered | | Reference | | 14.0% |
| | Offered | 0.19 (0.04) | 65.7 (59.0 to 72.0) | <0.001 | |

Log likelihood =-3493.69, Likelihood Ratio Test=247.24, Akaike's Information Criterion=7021.24

Table 9 (C). Participants with the most positive vaccine sentiments (n=334).

| Attribute | Level | Mean (standard error) | % who prefer this level (95% CI) | *p*-level | Relative importance |
|---|---|---|---|---|---|
| Vaccine | MMR | | Reference | | 15.0% |
| | 6-in-1 | 0.31 (0.07) | 63.4 (57.9 to 68.7) | <0.001 | |
| Child age group | 2 years and older | | Reference | | 17.0% |
| | 5 years and older | -0.35 (0.07) | 36.3 (31.3 to 41.5) | <0.001 | |
| Incentive | No incentive | | Reference | | 24.8% |
| | Incentive for child (£130 voucher) | 0.45 (0.06) | 79.1 (72.2 to 84.9) | <0.001 | |
| | Incentive for parent (£130 cash pay-out) | 0.51 (0.07) | 74.8 (68.8 to 80.0) | <0.001 | |
| Penalty | £450 fine | | Reference | | 21.1% |
| | Child not able to attend school or childcare / day care until they are vaccinated | 0.43 (0.07) | 70.3 (64.4 to 75.8) | <0.001 | |
| | Parent not able to claim Child Benefits for child until they are vaccinated | 0.20 (0.07) | 58.9 (52.5 to 65.0) | 0.006 | |
| Ability to opt out | Medical exemption | | Reference | | 8.9% |
| | Medical and religious belief exemption | -0.18 (0.05) | 38.4 (32.2 to 45.0) | 0.001 | |
| Compensation scheme | Not offered | | Reference | | 13.3% |
| | Offered | 0.27 (0.06) | 64.6 (58.9 to 70.0) | <0.001 | |

Log likelihood =-3254.93, Likelihood Ratio Test=575.25, Akaike's Information Criterion=6543.87

**Supplementary Materials 4. Principal components analysis of psychological factors**

Figure 1. Scree plot from principal component analysis conducted on items measuring psychological factors.

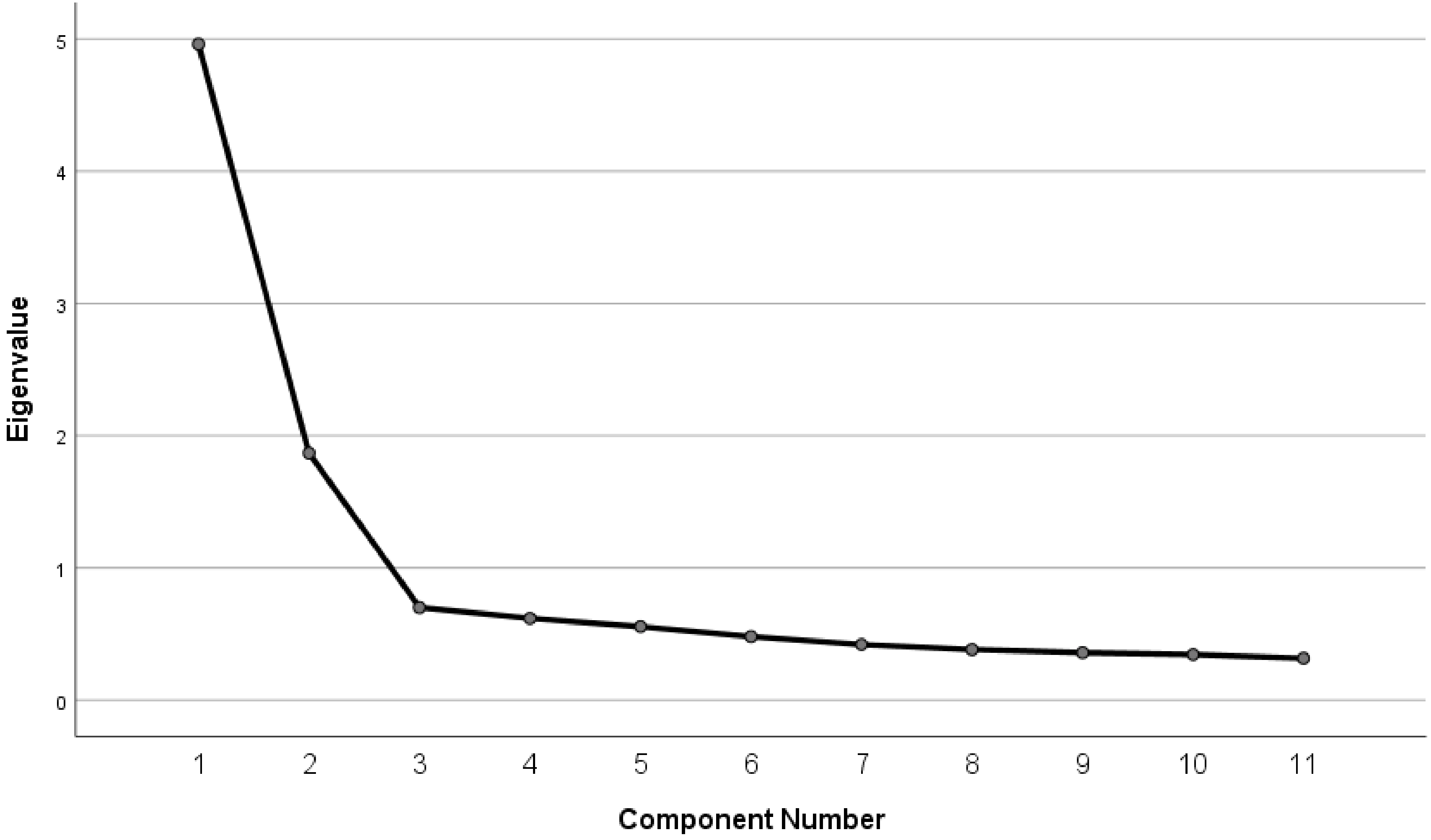

Table 1. Loadings of psychological factors onto components

| Component 1 – approval of child vaccines | | Component 2 – disapproval of child vaccines | |
|---|---|---|---|
| Item | Loading | Item | Loading |
| If I did not vaccinate [CHILD], [she/he] would be likely to catch the illnesses the vaccines aim to prevent | .81 | One of my children has had a severe side effect from a routine vaccination | .84 |
| If I did not vaccinate [CHILD], [she/he] could get severely ill from the illnesses the vaccines aim to prevent | .78 | Child vaccinations cause severe side effects | .82 |
| I approve of mandatory vaccination | .76 | Vaccination campaigns are just about making money for the manufacturers | .81 |
| Child vaccinations are an effective way of preventing children from catching vaccine-preventable illnesses | .73 | I don't like child vaccinations in general | .75 |
| If some children do not receive vaccinations, this may cause other children to be ill with the disease | .72 | Natural exposure to viruses and germs gives children the safest protection | .70 |
| Child vaccinations are safe | .70 | | |

Extraction method: principal component analysis. Rotation method: oblimin with Kaiser normalization. Rotation converged in 5 iterations.
